# Insights on Genetic and Environmental Factors in Parkinson’s Disease from a regional Swedish Case-Control Cohort

**DOI:** 10.1101/2021.06.18.21259024

**Authors:** Kajsa Brolin, Sara Bandres-Ciga, Cornelis Blauwendraat, Håkan Widner, Per Odin, Oskar Hansson, Andreas Puschmann, Maria Swanberg

## Abstract

**BACKGROUND:** Risk factors for Parkinson’s disease (PD) can be more or less relevant to a population due to population-specific genetic architecture, local lifestyle habits, and environmental exposures. Therefore, it is essential to study PD at a local, regional, and continental scale in order to increase the knowledge on disease etiology.

**OBJECTIVE:** We aimed to investigate the contribution of genetic and environmental factors to PD in a new Swedish case-control cohort.

**METHODS:** PD patients (n=929) and matched population-based controls (n=935) from the southernmost county in Sweden were included in the cohort. Information on environmental exposures was obtained using questionnaires at inclusion. Genetic analyses included a genome-wide association study (GWAS), haplotype assessment, and a risk profile analysis using cumulative genetic risk scores.

**RESULTS:** The cohort is a representative PD case-control cohort (64% men, mean age at diagnosis=67 years, median Hoehn and Yahr score=2.0), in which previously reported associations between PD and environmental factors, such as tobacco, could be confirmed. We describe the first GWAS of PD solely composed of PD patients from Sweden, and confirm associations to well-established risk alleles in *SNCA*. In addition, we nominate an unconfirmed and potentially population-specific genome-wide significant association in the *PLPP4* locus (rs12771445).

**CONCLUSIONS:** This work provides an in-depth description of a new PD case-control cohort from southern Sweden, giving insights into environmental and genetic risk factors of PD in the Swedish population.

## Introduction

Parkinson’s disease (PD) has been reported to be the fastest growing neurological disorder worldwide in terms of prevalence, disability, and deaths [1, 2]. Currently available treatments only address symptoms, and the burden of PD will increase substantially if no disease-modifying therapeutic is developed.

PD can be categorized into monogenic and idiopathic (multifactorial) based on the contribution of genetics to disease risk. In monogenic PD, rare DNA variants with large effect sizes but varying penetrance cause the disease [3]. Although such variants in *SNCA*, *LRRK2*, *VPS35*, *PRKN*, *PINK1, GBA,* and *DJ-1* [4–9] contribute to a small proportion of cases, these genes have provided valuable insight into the molecular mechanisms underlying PD etiology and pathogenesis, as well as an overlapping etiology between monogenic and idiopathic PD. Missense mutations and copy number variants (CNVs) of *SNCA*, encoding a-synuclein, have been identified to cause monogenic PD, whereas common *SNCA* variants have been associated with an increased risk of idiopathic PD [5]. In idiopathic PD, accounting for more than 95% of all PD cases, variants with relatively small effect sizes in combination with environmental exposures influence an individual’s disease risk [10, 11]. The latest large-scale genome-wide association study (GWAS) meta-analysis identified 90 risk loci associated with PD in individuals of European ancestry [12]. These variants were estimated to explain 16-36% of the heritable PD risk, indicating that many genetic risk factors for PD remain to be discovered [12]. Additionally, two new risk loci have been reported to be associated with PD risk in the Asian population [13].

Larger sample sizes in GWAS could pave the way for the discovery of additional risk variants and increase the proportion of explained heritability [14, 15]. However, it has also been suggested that the “missing heritability” could be explained by the presence of gene-gene and gene-environment interactions [14]. Commonly reported environmental risk factors for PD include smoking and exposure to pesticides, two exposures that can vary a lot between population. Further understanding environmental risk factors for PD in different populations are hence of great importance to further elucidate the pathogenesis of the disease [10, 16, 17].

There is a strong population bias towards individuals of European ancestry in genetic research of human diseases [18, 19], but initiatives are taken to study PD genetic risk in other populations [20, 21]. In addition to the need of a widened scope of PD genetic research on a global level, narrowed studies are also of interest, since heterogeneity across cohorts from different countries or regions may mask genetic associations specific to sub-population [22].

The Swedish population has not yet been represented in PD GWAS meta-analyses or population-specific GWAS aside from a PD GWAS performed on a mixed Scandinavian population from Norway and Sweden [23]. The Swedish population has been reported to be genetically similar to the nearby population of Norway [24, 25]. However, it has also been reported that Swedish genomes contain a substantial amount of genetic variation and genetic variants that are not represented in other European populations [26], and that there is a large genetic difference even within the Swedish population, particularly between the south and north of Sweden [24, 26]. This highlights the importance of analyzing different European populations, and emphasize the importance of regional matching of cases and controls in GWAS [24, 27].

Here, we provide a comprehensive description of a novel, well-defined, and matched PD case-control cohort from southern Sweden in which we describe, to our knowledge, the first GWAS of PD composed solely of individuals from Sweden. This study provides insights into environmental and genetic risk factors of PD in the Swedish population and will contribute to the understanding of population-specific environmental and genetic risk in PD.

## Materials and Methods

### Ethical consent

Ethical permission for the MultiPark’s biobank sample collection (MPBC) was approved by the regional ethics review board of Lund (2013/509). All participants gave informed written consent at study enrollment.

### Patient and population-based control inclusion

Individuals primarily diagnosed with PD (*International Classification of Disease*, *tenth revision (ICD-10-SE)* code G20.9) in the southernmost province of Sweden, Scania, were assessed for eligibility (Figure S1). Inclusion criteria were a PD diagnosis and the ability to visit one of the clinics for sampling. In total, 2,119 PD patients were invited to participate between November 2014 and July 2017, whereof 1,011 were included (inclusion rate 48.3%). For each patient, population-based controls matched by date of birth, sex, and residential area were randomly selected from the Swedish Population Register and invited between March 2015 and April 2018. A total of 1,001 individuals consented and completed data collection (inclusion rate 18.5%). Exclusion criteria were a PD diagnosis and the inability to visit a center for sampling. Out of the completed controls, 953 were unique matchings, resulting in a matched control for 953 PD patients (94.3%). All analyses were performed in a set of 929 patients and 935 controls passing all quality control (QC) steps unless stated otherwise. A flowchart detailing the study participant inclusion process can be found in Figure S2.

### Sample and data collection, storage, and management

Blood samples were handled by the Clinical Chemistry Unit at the University Hospital in Scania (SUS) and stored at -80°C within a central biobanking facility (Biobank Syd: BD47). Whole blood was collected in EDTA-tubes (Becton Dickinson (BD) Vacutainer®, New Jersey, US) for DNA extraction. Additional samples were collected in PAXgene blood RNA tubes (QIAGEN®, Hilden, Germany), EDTA-tubes (BD Vacutainer®), sodium-heparin tubes (BD Vacutainer®, New Jersey, US) and a serum-separating tube (BD Vacutainer® SST™, New Jersey, US) to enable future studies of RNA and biomarkers in PD. When difficulties emerged during blood sample collection, EDTA-tubes for DNA extraction were prioritized. Serum and plasma samples are stored at -80°C in a 96-well format of 200 µl aliquots.

All participants filled in questionnaires covering basic demographic data, lifestyle habits, family history of PD, comorbid diseases, environmental exposures, medications, health status, and perceived motor- and non-motor symptoms. Data from questionnaires was manually entered into the web-based application REDCap [28]. Additional data for the PD rating tools Hoehn and Yahr (H&Y), Clinical Impression of Severity Index - Parkinson’s Disease (CISI-PD), and Parkinson’s Disease Questionnaire - 8 (PDQ-8), as well as information on age at diagnosis (AAD) was retrieved from the Swedish Parkinson’s registry (https://www.neuroreg.se/parkinsons-sjukdom/).

### Statistical analyses of epidemiological data

Epidemiologic data was analyzed using R version 4.0.0 [29]. Demographic characteristics were summarized as frequency and percentage for categorical variables. For continuous variables, mean and standard deviation (SD) were used for variables displaying normal distribution whereas median and interquartile range was reported for non-normally distributed variables. The range for continuous variables was used to demonstrate the heterogeneity in the cohort. Body mass index (BMI) was calculated as weight (kg)/height (m)^2^. For quality-of-life assessment, EuroQol five-dimension-3-level (EQ-5D-3L) was used, covering mobility, self-care, usual activities, pain/discomfort, and anxiety/depression. Time trade-off (TTO) and the visual analogue scale (VAS) index were calculated using scale value sets for Sweden [30]. In TTO valuation, individuals were asked to indicate how many years in full health that would be of equal value to 10 years in their current health state (divided by 10). Full health hence corresponds to 10 years/10=1. In VAS valuation, respondents rated their overall health between 0 (worst imaginable health) and 100 (best imaginable health) [31]. Additionally. the occurrence of PD symptoms at study inclusion were compared between the patients and controls using Pearson’s chi square test with a Bonferroni corrected threshold for statistical significance of p=0.001.

To test for associations between potential risk factors and PD status, both unadjusted and multivariate logistic regression analyses were used. Risk factors were selected based on meta-analysis literature reviews [16, 17]. Due to the cross-sectional case-control study design, we selected variables from the questionnaire related to past exposures, excluding variables related to exposures at time or near time of inclusion. Directed acyclic graphs (DAGs) were made for all exposures of interest using the R package ‘dagitty’ to help identify confounding variables to adjust for in the multivariate analyses (Figure S3) [32]. Confounding pathways with the least amount of missing data were prioritized and complete-case analyses were run. Odds ratios (OR), and 95% confidence intervals (CI) were estimated and plotted in a forest plot.

We further evaluated variables for BMI and comorbidities at inclusion, along with alcohol and red wine consumption and physical exercise the year prior to inclusion, and ibuprofen use the past two weeks prior to inclusion, using logistic regression analyses, adjusting for age and sex. We acknowledge the risk of confounding factors but the interest was solely to perform observational analyses. Complete-case analyses were run and ORs and 95% CI were estimated and plotted in a forest plot. A total of 35 comorbidities were not evaluated for association due to either too few study participants reporting a certain diagnosis or a lack of difference between the occurrence of the diagnosis between the patient and control group.

### DNA extraction and Genotyping

DNA extraction was done from whole blood at BD47 and at LGC Biosearch Technologies (UK, GEN-9300-120). LGC Biosearch Technologies (Germany) also performed genotyping and subsequent “Basic BioIT” with technical QC and genotype clustering. The no-call GenCall threshold was set to the standard score cutoff for Infinium data of 0.15. Matched case-control samples were genotyped using the Infinium Global Screening Array-24 v.2.0 with the Multi-Disease drop in panel v.2.0 (GSAMD-24v2.0) containing 712,189 variants annotated to the Genome Reference Consortium Human Build 37 (GRCh37).

### Genotype quality control and Imputation

QC analysis was performed in PLINK 1.9 [33, 34]. Samples were excluded if the call rate was <95%, if excess heterozygosity was detected (estimated by a F statistic >0.15 or <-0.15), or if the genetic sex (determined from X-chromosome heterogeneity) differed from the reported sex.

Principal component analysis (PCA) was used to identify and exclude ancestry outliers. For this, the genotype data was merged with the HapMap phase 3 data set [35], and samples were determined as having European ancestry if they clustered 6 SD (+/-) around the combined population mean of the populations: Utah residents with Northern and Western European ancestry (CEU) and Toscani in Italia (TSI) (Figure S4). Samples from closely related individuals, defined as sharing more than 12.5% of alleles, were also excluded. Following sample exclusion, 929 patients and 935 controls remained. Variant level QC was subsequently done, excluding SNPs with a missingness rate >5%, Hardy-Weinberg equilibrium p-value <1E-4, or minor allele frequency (MAF) <1%. SNPs with differences in genotyping rate for patients and controls, and genotypes missing by haplotype, were also excluded for p-value <1E-4. SNPs passing QC were 505,005 with a total genotyping rate of 99.9%. Genomic imputation was performed on the Michigan Imputation Server [36] using the Minimac4 imputation software, Eagle v2.4 for phasing [37], and the Haplotype Reference Consortium (HRC) r1.1 2016 European population as the reference population (http://www.haplotype-reference-consortium.org/) [38]. Post-imputation QC was performed, and variants with MAF<5% and/or poor imputation quality, set as Rsq <0.3, were excluded. The number of imputed variants after filtering was 5,445,841.

### Power calculation for GWAS analyses

GWAS power calculation was performed using the online Genetic Association Study (GAS) Power Calculator (http://csg.sph.umich.edu/abecasis/gas_power_calculator/). In order to reach 80% power at a GWAS-significance level (alpha=5E-08) with a disease prevalence in the general Swedish population of 0.2%, an allele frequency of >20% and an OR>1.7 would be needed.

### Genome wide association study (GWAS) vs PD risk and age at diagnosis (AAD)

A GWAS for PD risk was performed in a logistic regression model adjusted for sex, age (at study inclusion), and the first 5 principal components (PCs). The number of PCs used in the analyses was determined by a scree plot (Figure S5). A linear regression model was used to investigate the impact of genetic variation on the AAD, adjusted for sex and PC1-5. Data on AAD was available for 792 of the 929 PD patients (85.3%). Summary statistics was generated using RVTESTS (version 2.1.0) [39]. Quantile-quantile (QQ)-plots and Manhattan plots were generated in R, version 4.0.0 [29]. Subsequently, regions of interest were visualized using summary statistics data in the LocalZoom tool (https://my.locuszoom.org) with a flanking size of 100 kb from the gene of interest.

### Haplotype analysis of *PLPP4*

Genotyped variants in the *PLPP4* region with a window of ±100 kb (*PLPP4* coordinates on GRCh37; chr10:122216466-122349376, UCSC) was extracted and annotated using ANNOVAR [40]. Haplotype blocks in the region were estimated in PLINK v1.9 using the default parameters wherein pairwise LD was calculated for variants within 500 kb [33, 34]. PLINK v1.9 estimates haplotype blocks following the default procedure in Haploview [41, 42]. Association between haplotype blocks in *PLPP4* and PD status was analyzed with haplo.stats in R. The regression analyses were adjusted for age, sex, and PC1-5, and additionally for tobacco use. The adjustment for tobacco use was done because of a report of *PLPP4* being associated with smoking cessation [43], and multiple reports of smoking being associated with PD. To assimilate the effect of other nicotine products, such as snus (moist tobacco, commonly consumed in Sweden), the variable “Tobacco” was used. Only haplotypes with a carrier frequency of >1% were included in the analysis. A Bonferroni corrected threshold for statistical significance of p=0.05/16=0.003. was set. The results were visualized using LocusZoom (https://my.locuszoom.org) and Haploview v4.1 [44].

### Analysis of the joint effect of variants in *PLPP4* on PD risk

The contribution of rare variants on PD risk was examined by the joint effect of multiple variants in the *PLPP4* region. We separately analyzed genotyped and imputed variants with an imputation quality score of Rsq >0.8 for imputed variants. The sequence kernel association test (SKAT) was used since only 2.6% of the variants in *PLPP4* are coding variants and the SKAT is powerful when a large fraction of variants in a region are noncoding. The test aggregates associations between variants and phenotype and allows for variant interactions [45]. The analyses were performed in RVTESTS (version 2.1.0) [39] using default parameters and adjusting for age, sex and PC1-5. The test was performed both for all available variants as well as in only coding variants with two different maximum MAF thresholds of <1% and <5%. To adjust for multiple comparisons, Bonferroni corrected p-value thresholds was applied for all analyses.

### Risk profile analysis

A risk profile analysis for PD and AAD was performed using the imputed data passing QC. A cumulative genetic risk score (GRS) was calculated in PLINK v1.9 using publicly available effect estimates (beta coefficients) of 90 SNPs associated with PD in the largest published meta-analysis of PD GWAS to date [12]. For each SNPs, the allele dosage was multiplied by the beta coefficient, giving a greater weight to alleles with higher risk estimates. Risk allele dosage was then summed across all variants to generate a GRS for each study participant. Subsequently, the GRSs were standardized to Z-scores using the control group as the reference group for PD status and the patient group for the AAD analysis. Following standardization, a Z score of 1 can be interpreted to be equivalent to one SD increase in the GRS from the reference group GRS mean. The study participants were divided into quartiles based on their GRS, and logistic regression analysis was performed to investigate associations and estimate the OR between PD status and quartile group. Covariates were selected from a previously published PD classification [46] and included PD family history (yes/no), age at inclusion, sex, and additionally PC1-5. For the risk profile analysis for AAD, a linear regression model was used, adjusted for sex, PD family history, and PC1-5. Plots were generated in R, version 4.0.0 [29].

Code for all analyses will be available at https://github.com/KajBro/MPBC, GWAS summary statistics will be available online.

## Results

### Characteristics of the MPBC cohort

Demographics and characteristics of the MPBC cohort are summarized in Table 1. An expected male predominance was observed with a sex ratio of 2:1. Case-control matching was successful for year of birth and sex, with average birth year 1944 and 36% women in both groups. A one-year difference for the average age at inclusion was observed between patients (71 years) and controls (72 years) due to control matching occurring after patient inclusion. Swedish ancestry (defined as self-reporting that both parents were born in Sweden) was similar in patients and controls (87.9% vs 86.1%). Similar distributions for highest completed education and marital status were also seen. According to self-reported information on health status using the EQ-5D-3L instrument [30], patients had an index of 0.8 on the TTO scale compared to 0.9 for the controls. The patients also rated their own overall health lower than the controls on the VAS index scale (index 68.9±16.8 vs 82.0±9.7, Table 1). As a reference, the general Swedish population has an estimated mean TTO index of 0.9 and a VAS index of 79.5 [30]. As expected, all PD symptoms were more common in the patient group than the control group (Table S3). The most common self-reported motor-symptoms were muscle stiffness (72.7%) followed by slowness of movements (72.4%). The most commonly reported non-motor symptom for both patients and controls was nocturia (71.6% vs 58.4%).

**Table 1:** Characteristics of participants (individuals with Parkinson’s disease (PD) and individuals without the disease [controls]) in the MPBC cohort.

|  | Patients (N=929) | Controls (N=935) |
| --- | --- | --- |
| **Sex** |  |  |
| n | 929 | 935 |
| Men | 599 (64.5%) | 598 (64.0%) |
| Women | 330 (35.5%) | 337 (36.0%) |
| **Birth year** |  |  |
| n | 929 | 935 |
| Mean (SD) | 1944.4 (8.4) | 1944.4 (8.3) |
| Range | 1920.0-1979.0 | 1920.0-1980.0 |
| **Age at inclusion** |  |  |
| n | 929 | 935 |
| Mean (SD) | 71.0 (8.3) | 72.3 (8.3) |
| Range | 37.0-96.0 | 38.0-97.0 |
| **Swedish ancestry** |  |  |
| n | 922 | 921 |
| No | 112 (12.1%) | 128 (13.9%) |
| Yes | 810 (87.9%) | 793 (86.1%) |
| **Highest completed education** |  |  |
| n | 923 | 929 |
| < Primary and lower secondary education | 8 (0.9%) | 4 (0.4%) |
| Primary and lower secondary education | 241 (26.1%) | 232 (25.0%) |
| Upper secondary education | 403 (43.7%) | 384 (41.3%) |
| University | 271 (29.4%) | 309 (33.3%) |
| **BMI at inclusion (kg/m<sup>2</sup>)** |  |  |
| n | 914 | 921 |
| Mean (SD) | 25.8 (4.4) | 26.0 (3.9) |
| Range | 15.2-48.8 | 15.9-42.3 |
| **Marital status** |  |  |
| n | 925 | 931 |
| Single | 57 (6.2%) | 62 (6.7%) |
| Married/Civil partnership | 719 (77.7%) | 713 (76.6%) |
| Divorced/Separated | 71 (7.7%) | 60 (6.4%) |
| Widow/Widower | 78 (8.4%) | 96 (10.3%) |
| **TTO Index** |  |  |
| n | 898 | 903 |
| Mean (SD) | 0.8 (0.1) | 0.9 (0.1) |
| Range | 0.4-1.0 | 0.5-1.0 |
| **VAS Index** |  |  |
| n | 898 | 903 |
| Mean (SD) | 68.9 (16.8) | 82.0 (9.7) |
| Range | 17.2-88.9 | 20.9-88.9 |

Swedish ancestry: Both parents were born in Sweden. BMI: Body Mass Index; TTO: Time Trade Of, EQ-5D-3L;
VAS: Visual Analogue Scale, EQ-5D-3L

We further evaluated the health status and AAD among male and female PD patients (Table 2). Median AAD was identical for both sexes (67.0 years), although with a wide range (men 29.0-84.0 years, women 35.0-89.0 years). The median time since diagnosis at inclusion was 4.0 years for both sexes and varied from diagnosis the same year to 36.0 years for men and 33.0 years for women. Health status at the time of inclusion was evaluated using the H&Y scale, CISI-PD, and PDQ-8. Both sexes had a median of 2.0 on the H&Y, corresponding to bilateral involvement without impairment of balance. Similar total score values were reported by men and women on the CISI-PD scale (median 5.0, range 0-24). Women had a slightly higher median PDQ-8 total score compared to men (7.0 vs 6.0 out of 32).

**Table 2:**
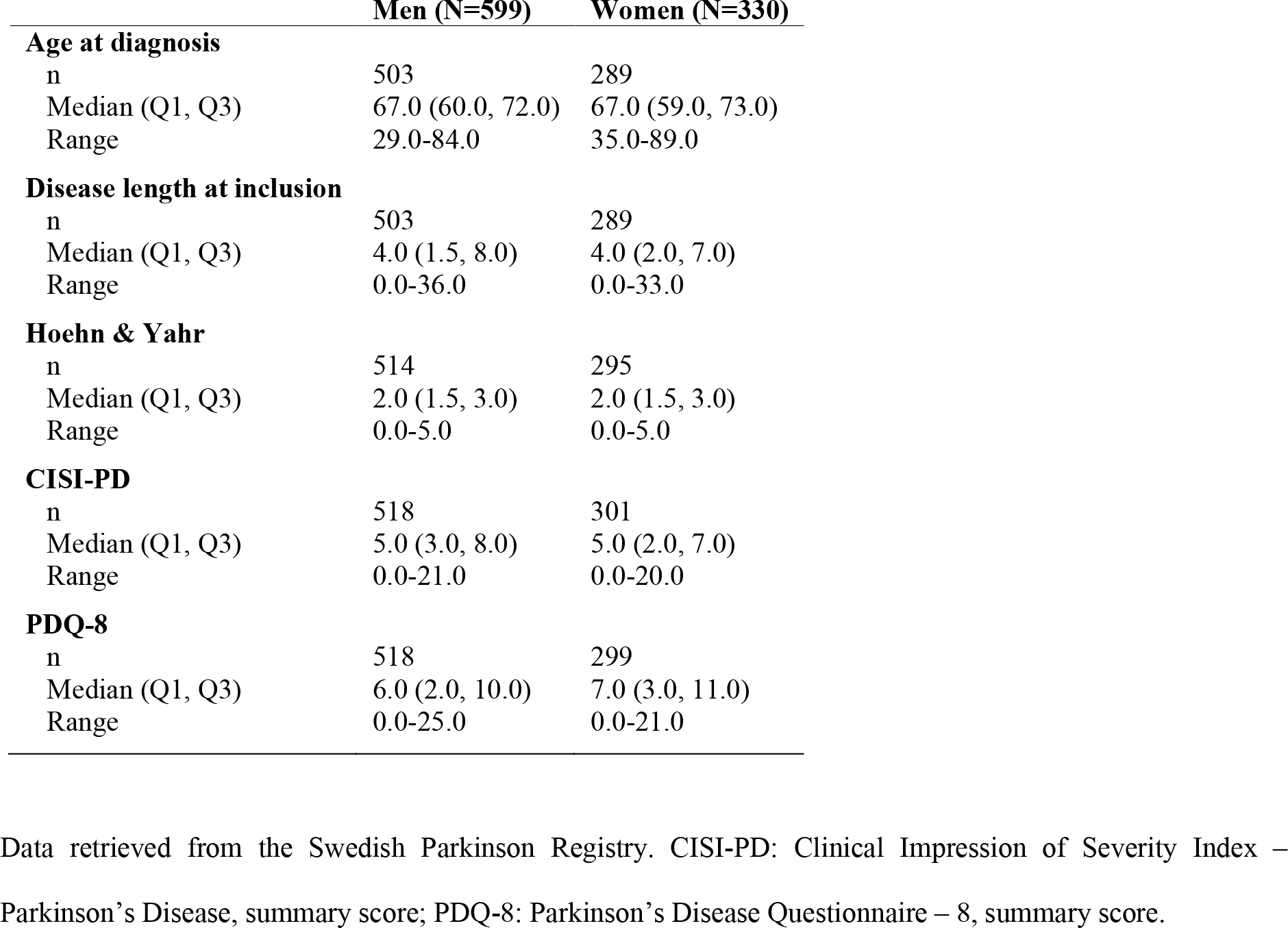
Characteristics of individuals with Parkinson’s disease in the MPBC cohort.

|  | **Men (N=599)** | **Women (N=330)** |
| --- | --- | --- |
| **Age at diagnosis** |  |  |
| n | 503 | 289 |
| Median (Q1, Q3) | 67.0 (60.0, 72.0) | 67.0 (59.0, 73.0) |
| Range | 29.0-84.0 | 35.0-89.0 |
| **Disease length at inclusion** |  |  |
| n | 503 | 289 |
| Median (Q1, Q3) | 4.0 (1.5, 8.0) | 4.0 (2.0, 7.0) |
| Range | 0.0-36.0 | 0.0-33.0 |
| **Hoehn & Yahr** |  |  |
| n | 514 | 295 |
| Median (Q1, Q3) | 2.0 (1.5, 3.0) | 2.0 (1.5, 3.0) |
| Range | 0.0-5.0 | 0.0-5.0 |
| **CISI-PD** |  |  |
| n | 518 | 301 |
| Median (Q1, Q3) | 5.0 (3.0, 8.0) | 5.0 (2.0, 7.0) |
| Range | 0.0-21.0 | 0.0-20.0 |
| **PDQ-8** |  |  |
| n | 518 | 299 |
| Median (Q1, Q3) | 6.0 (2.0, 10.0) | 7.0 (3.0, 11.0) |
| Range | 0.0-25.0 | 0.0-21.0 |

Data retrieved from the Swedish Parkinson Registry. CISI-PD: Clinical Impression of Severity Index – Parkinson's Disease, summary score; PDQ-8: Parkinson's Disease Questionnaire – 8, summary score.

### Epidemiological characteristics of PD in Sweden

We further set out to evaluate previously confirmed risk factors for PD in a regional, Swedish context (Figure 1, Table S1). A positive family history (at least one relative diagnosed with PD) was overrepresented among PD patients compared to controls (20% vs 11%). Consequently, having a relative diagnosed with PD doubled the risk of having PD (OR=2.00, 95% CI 1.51-2.67). An association between pesticide exposure and PD was confirmed at an OR of 2.26 (95% CI 1.39-3.72). A significant association was also seen between PD and a history of head trauma (OR=1.30, 95% CI 1.08-1.58). However, no association between loss of consciousness and PD was observed among study participants who reported a history of head trauma. A slightly higher OR was seen for a higher BMI at the age of 20 years (1.05, 95% CI 1.01-1.09), indicating that a one unit increase in BMI increased the risk of having PD by 5%. However, no difference was observed for the highest reported BMI. We observed an inverse association between having ever smoked and PD, but the association was not statistically significant after adjustment for confounders (ever-vs never-smoking OR=0.82, 95% CI 0.67-1.01). A statistically significant inverse association between snus and PD was observed also after adjusting for confounders (OR=0.53, 95% CI 0.38-0.73). An inverse association was also observed for the combined effect of tobacco products on PD (adjusted OR=0.72, 95% CI 0.59-0.88). Moreover, a trend of lower OR for PD with increasing amount of coffee drinking at all investigated age groups was observed, where drinking >5 cups of coffee per day was inversely associated with PD (OR 0.36-0.52, 95% CI 0.18-0.86).

**Figure 1:**
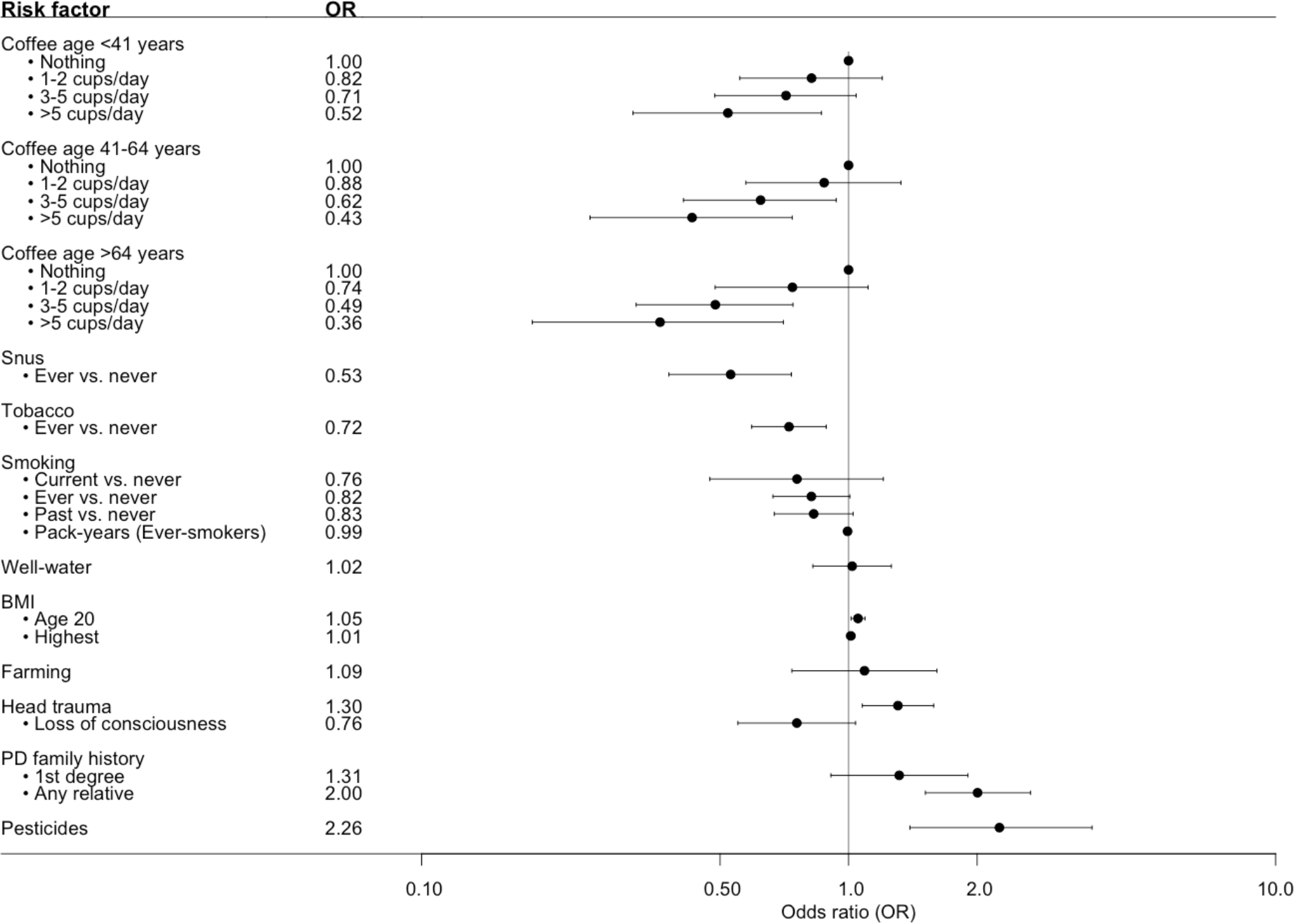
Forest plot over the associations between risk factors and PD in MPBC. Showing the adjusted OR and 95% CI. Number of individuals in each analysis is found in Table S1.

Additionally, variables related to exposure/use within the past year were examined to describe potential differences between the age- and sex-matched groups at inclusion (Figure 2, Table S2; Table S3). The patients consumed less alcohol and engaged in less physical activity both in regard to level of physical activity (regular activity level vs nothing OR=0.26, 95% CI 0.18-0.38) and hours per week (≥5 hours/week vs nothing, OR=0.21, 95% CI 0.13-0.33). No significant association between PD and BMI at inclusion was observed (OR=0.99, 95% CI 0.96-1.00). For comorbidities, statistically significant inverse associations between PD and a diagnosis of hyperlipidemia (OR=0.51, 95% CI 0.40-0.64), hypertension (OR=0.57, 95% CI 0.47-0.69), and osteoarthritis (OR=0.66, 95% CI 0.53-0.82) were seen. A significant association with PD was observed for a diagnosis of depression (OR=1.89, 95% CI 1.39-2.59), back pain (OR=1.56, 95% CI 1.15-2.13), and bowel problems, where bowel problems (defined as having constipation or diarrhea that required treatment on a regular basis) almost quadrupled the likelihood of having PD (OR=3.93, 95% CI 2.66-5.97). We observed an OR of 0.53 (95% CI 0.41-0.68) for the use of ibuprofen <2 times/week compared to never. However, the association was lost for ibuprofen use ≥2 times/week. No associations were observed for the use of other nonsteroidal anti-inflammatory drugs (NSAIDs), such as diclofenac or naproxen (data not shown).

**Figure 2:**
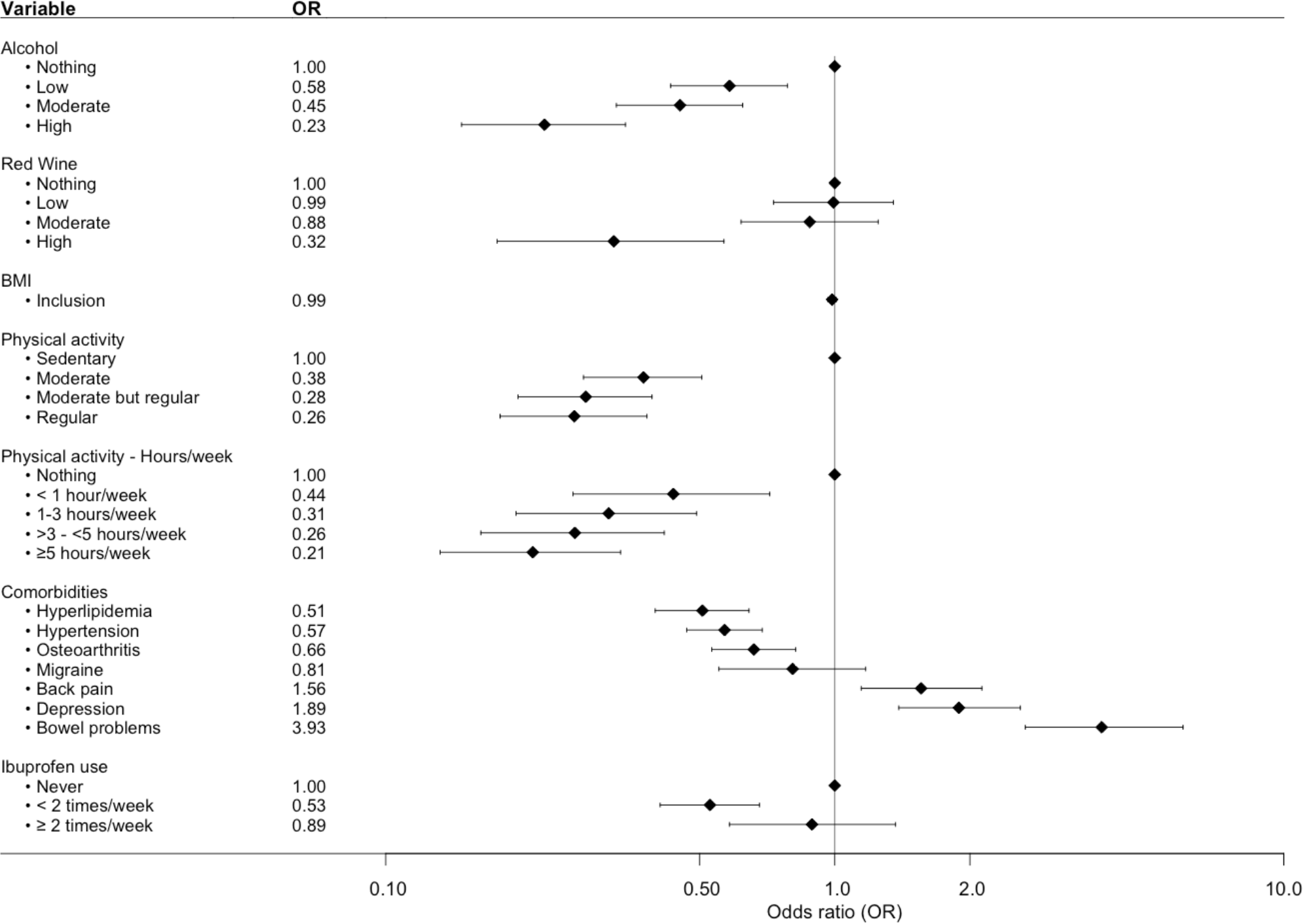
Forest plot over the associations between various variables and PD. Analyzed variables were obtained from questions regarding exposure/use within the past year prior to study inclusion. The plot shows OR and 95% CI adjusted by age at inclusion and sex. Number of individuals in each analysis is found in Table S2.

### GWAS of PD risk nominated a novel genome-wide significant variant in the *PLPP4* locus

The variant rs356182 in *SNCA* was reported in the latest GWAS meta-analysis as one of the 90 risk loci associated with PD in a cohort of European ancestry [12]. In our GWA analysis for PD risk, this and another three variants in the *SNCA* locus were observed at low p-values (rs356182, beta=-0.38, SE=0.07, p=5.64E-08; rs356203, beta=-0.36, SE=0.07, p=1.27E-07; rs356220, beta=0.36, SE=0.07, p=1.39E-07; rs356219, beta=-0.36, SE=0.07, p=2.32E-07), but none reached genome-wide significance (Table S4). In addition to the *SNCA* variants, we found associations for 23 of the 90 previously reported PD risk loci in this Swedish cohort at an uncorrected p-value of 0.05 (Table S5). Results from PCA showing the population structure as compared to the HapMap3 reference populations, revealed that this Swedish cohort cluster near the other European population but the centers of the clusters differ slightly, indicating differences in population structures that could result in population-specific risk alleles (Figure S6). The GWA analysis also revealed a novel genome-wide significant association signal in the *PLPP4* locus, rs12771445 (10:122318147, beta=-0.44, SE=0.07, p=1.30E-08), as being associated with PD in this Swedish cohort (Figure 3, Table S4). This association has, to our knowledge, not been reported in any other cohort (Table S4) [12]. The significant variant was an intron variant in the gene Phospholipid Phosphatase 4 (*PLPP4*) and was imputed with Rsq=0.99 and MAF=0.31 (Table S4). The MAF is similar to the value observed in the Swedish reference population, SweGen, of 0.32 [26]. The estimated MAF was 0.27 in the patient group and 0.36 in the control group, indicating a potential protective effect of the minor allele (T) with an OR of 0.64. To further display the association signals relative to genomic position and local LD, locus zoom plots for *PLPP4* rs12771445 and *SNCA* rs356182 were produced (Figure 4 and Figure S10). Variants in *SNCA* have been reported to also be associated with PD AAO [47]. We performed a GWA for the related variable AAD of PD in the cohort. However, we did not detect any genome-wide statistically significant associations and could not replicate previous reported variants associated with PD AAO at low p-values, likely due to smaller sample size (792 vs 28,568) [47], and limited statistical power (Figure S11 and S12).

**Figure 3:**
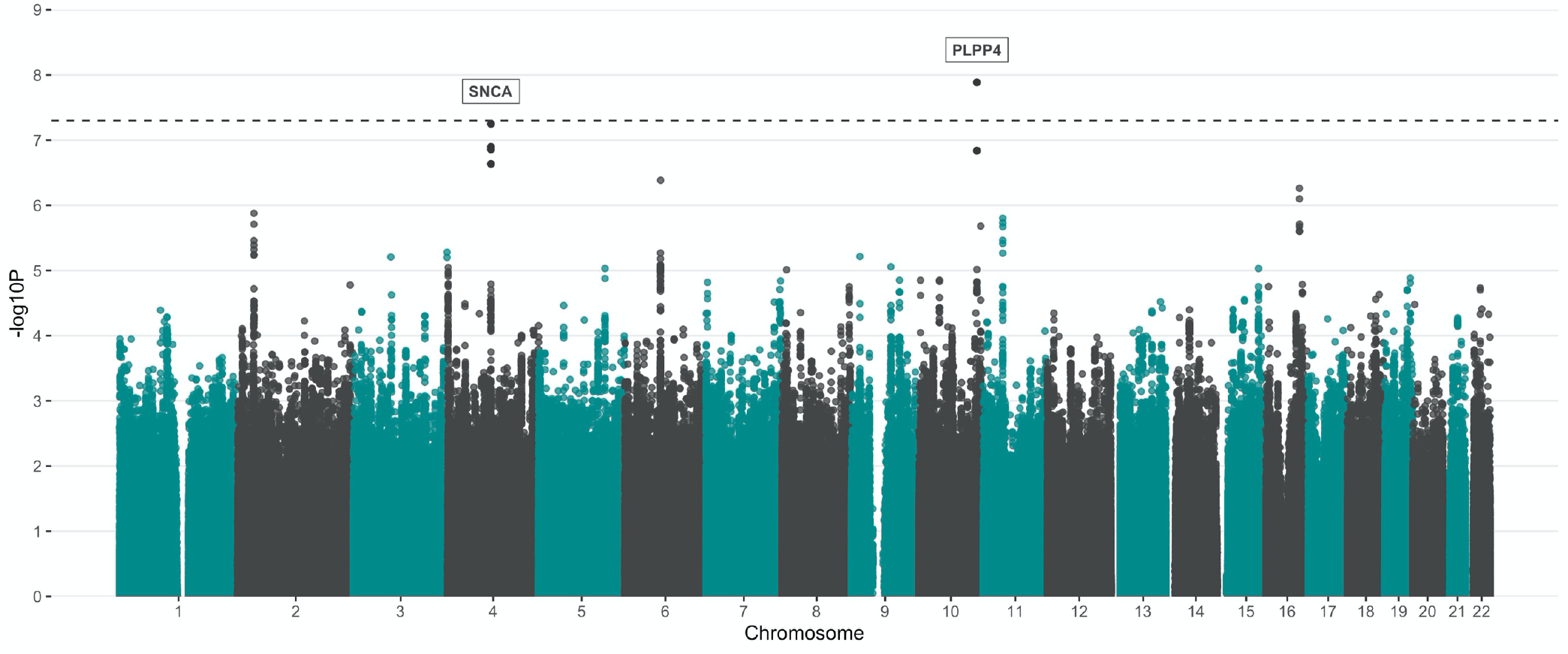
Manhattan plot showing the result from PD GWA analysis. A total of 5,445,841 SNPs (MAF >5%) were tested for 929 PD patients vs 935 controls. The y-axis represents the negative log (two-sided p-values) for association of variants with PD and the x-axis represent the genomic position on genome build GRCh37. The horizontal dashed line indicates the genome-wide significance level (p=5E-08).

**Figure 4:**
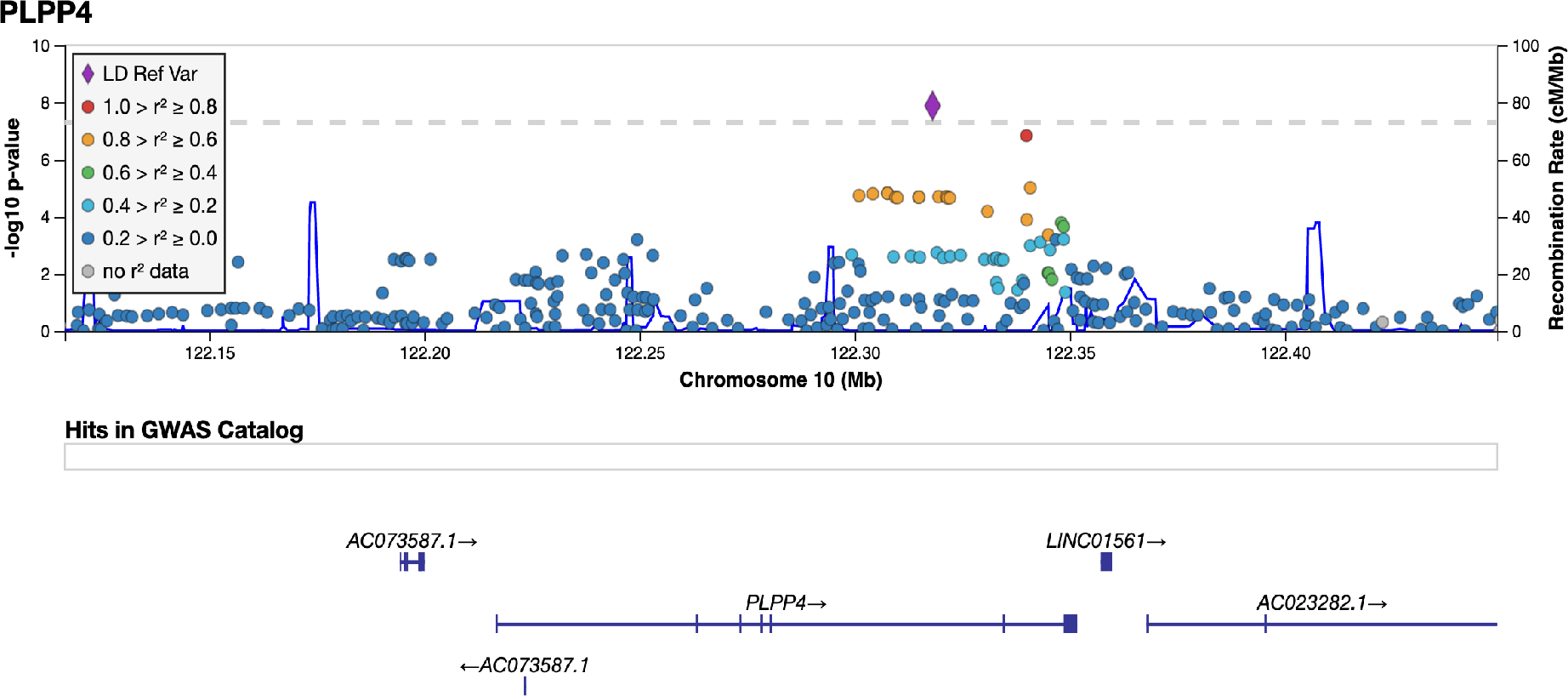
LocusZoom plot for PD GWAS PLPP4 loci. Imputed and genotyped variants passing QC in the *PLPP4* gene +/- 100 kb (10:122116466-122449376) mapped to genome build GRCh37. The only coding gene in the region is *PLPP4*, other include pseudogenes (AC073587.1) and long non-coding RNA (LINC01561, AC023282.1, WDR11-AS1). The variant with lowest p-value (index) is indicated as a purple diamond. Marker colors indicate the strength of LD as r^2 between the index variant and other variants in the 1000 Genomes EUR population.

In order to analyze more genetic variants, imputation was done using the beta-version of the TOPMed Imputation Reference panel [48]. Using the same post-imputation QC, 6,214,098 variants were generated and analyzed in a GWAS for PD risk (Figure S8 and S9). Two variants passed genome-wide significance, rs12772937 (chr10:122318146, beta=-0.40, SE=0.07, p=2.48E-08) and our previously identified rs12771445 (10:122318147, beta=-0.40, SE=0.07, p=2.48E-08). Since the TOPMed panel was still in the beta stage as of our analysis, we continued using the data imputed with the HRC version r1.1 2016 panel.

### Haplotype analysis of *PLPP4*

Because of the identification of the novel genome-wide signal (rs12771445) associated with PD in this cohort, we further looked at haplotypes among the genotyped variants in the nearest gene, *PLPP4* (± 100 kb), to capture the genetic variance in the region. We identified 92 genotyped variants passing QC in the region, where 32 were in *PLPP4*, all intron variants. The median GenCall scores of these variants was 0.87 (IQR: 0.83–0.91) (Figure S13). A total of 45 different haplotypes in 12 different blocks (H1-H12) in the region was identified (Table 3, Figure S14). Statistically significant association with PD was observed for a haplotype in one block (H6) (p=2.45E-05), both before and after adjustment. The haplotype block spanned over 41.9 kB (chr10:122302038-122343950) and consisted of five genetic variants and six different haplotypes. The rs978854(G)-rs7910507(G)-rs10886711(G)-rs11199417(G)-rs10886717(A) haplotype was statistically significantly inversely associated with PD at an OR of 0.69 (95% CI 0.59-0.82, p=2.54E-05) and with a MAF=0.24 for patients and 0.30 for controls.

**Table 3:**
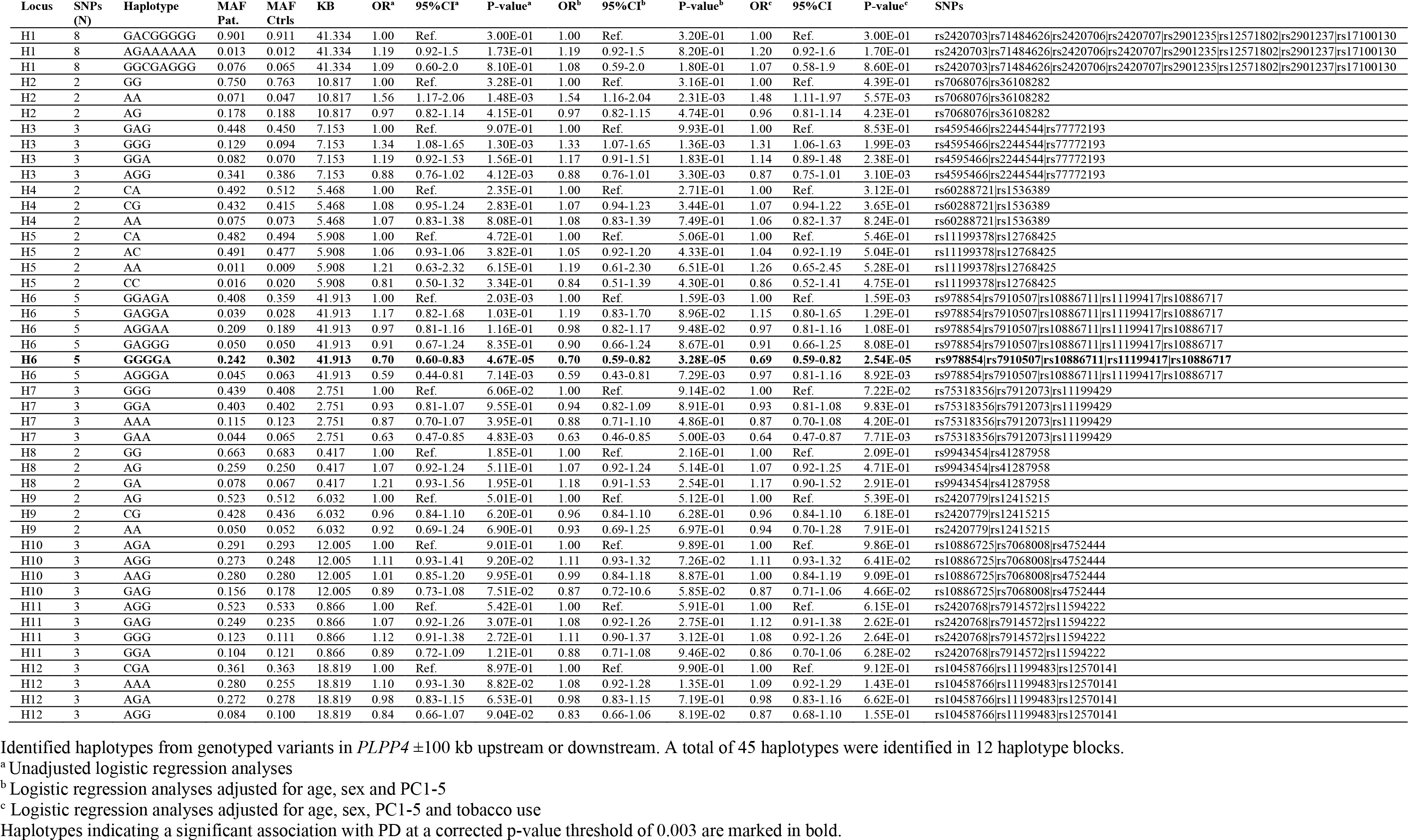
Identified haplotypes in the *PLPP4* locus.

**Table 4:** Number of variants in *PLPP4* tested through SKAT and the results statistics at MAF threshold 1% and 5% in the genotyped data vs the imputed.

|  | MAF < 1% |  | MAF < 5% |  |
| --- | --- | --- | --- | --- |
|  | No. Variants | p-value | No. Variants | p-value |
| **Genotyped data** |  |  |  |  |
| All variants | 1 | NA | **16** | **0.002** |
| Coding variants | 0 | NA | 0 | NA |
| **Imputed data** |  |  |  |  |
| All variants | 402 | 0.109 | 628 | 0.005 |
| Coding variants | 1 | 0.562 | 1 | 0.562 |

Analyses were adjusted for age at inclusion, sex and PC1-5 Results significant at Bonferroni correction are highlighted in bold

### Analysis of the joint effect of variants in *PLPP4* on PD risk

We further evaluated the joint effect of more rare variants (MAF <5%) in *PLPP4* on PD risk using both genotyped and imputed data. No coding variants in the gene were identified in the genotyped data and only one was identified in the imputed data (Table 5). When focusing on all variants, an association in the genotyped data at MAF<5% was observed (p=0.002), passing Bonferroni correction. A trend of a potential joint effect of variants with a MAF<5% was still observed in the imputed data (p=0.005). However, it did not reach multiple testing corrections (p-value threshold=0.05/628=7.96E-05). Only one variant with MAF<1% was present in the genotyped data and no statistically significant association was observed for variants with MAF<1% in either the genotyped or the imputed data.

### Genetic PD risk profile

The PD risk profile analysis showed that the Z-standardized GRS from Nalls et al was associated with PD status also in this Swedish case-control cohort [12]. For each SD increase from the reference mean value, the OR increased by 1.8 (beta=0.59, SE=0.05, p<2.22E-16) (Figure 5). Study participants in the highest GRS quartile were estimated to be 4.7 times more likely to have PD compared to the participants in the lowest quartile using an adjusted model. There was also an inverse association between the GRS and AAD, where one SD increase in the Z-standardized GRS was associated with approximately one-year earlier AAD (beta=-0.97, SE=0.36, p=0.007, adjusted r^2^=0.011).

**Figure 5:**
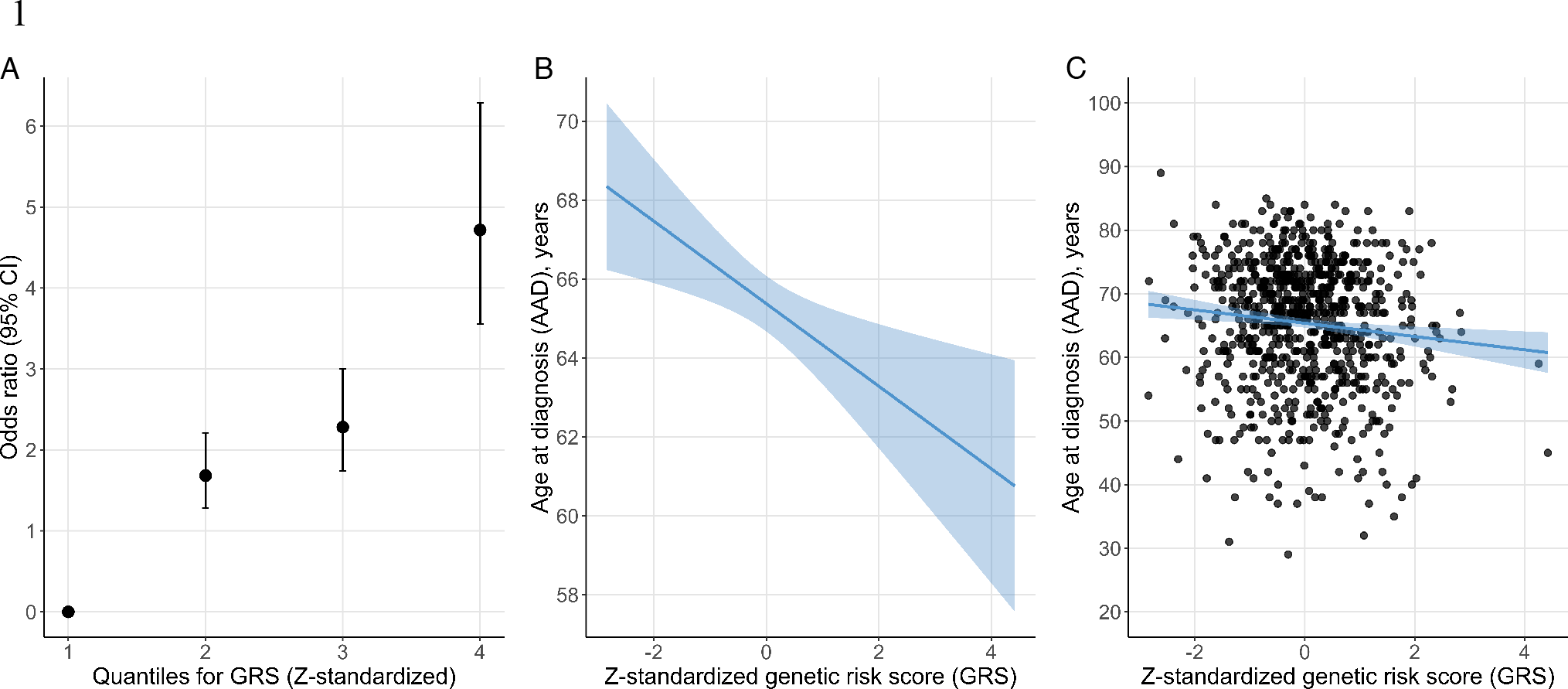
Genetic risk score (GRS) quartiles vs disease status (A) and age at diagnosis (AAD) (B and C). A: Odds ratio of PD status per risk quartile of the Z-standardized GRS. B: Regression line for the association between the Z-standardized GRS and AAD. The line represents the parameter estimate and the shading the 95% CI of the regression model. The model was adjusted for sex, PD family history, and PC1-5. C: AAD and Z-standardized GRS for each study participant with the regression model in plot B fitted to the plot.

## Discussion

This study is, to our knowledge, the largest case-control study of PD performed in a Swedish cohort. The study includes a well-defined recruitment of matched study participants from a specific region in southern Sweden. Multiple previously reported environmental and genetic risk factors were confirmed to affect PD risk. Interestingly, a novel genome-wide significant association with PD risk at the *PLPP4* locus was observed, both in the GWA and the haplotype analysis but further validation of the nominated variant is needed.

The MPBC cohort demonstrates similar characteristics compared to other PD cohorts. At inclusion, the patients had a relatively short disease duration (average 4 years), which was reflected in the PD rating scales with an average score of 2 on the H&Y scale and low scores on the CISI-PD and PDQ-8 scales. This can be explained by the study inclusion design where patients with advanced PD, who could not visit a neurology clinic, were not eligible. For self-reported PD symptoms, hypokinesia/bradykinesia was most common, occurring in 72% of the patients. Non-motor symptoms occurring at later stages of the disease, such as delusions and hallucinations were less frequent (6.0% and 16.8%), also highlighting the lower number of patients with advanced PD in the cohort [49]. The lack of an association between BMI at inclusion and PD further highlight this. Weight loss has frequently been observed in PD patients as a consequence of reduced energy intake in combination with an increased energy expenditure due e.g. levodopa-induced dyskinesia [50]. It has been estimated that PD patients lose 3.5 kg every eight years post-diagnosis and that levodopa-induced dyskinesia commonly develops after 3-5 years [50, 51]. Approximately 92% of the patients in the cohort reported using levodopa therapy.

Several previously reported environmental PD risk factors could be confirmed in this Swedish cohort. Among those, an association between pesticide exposure and PD was observe, where exposure to pesticides more than doubled the risk of having PD. The number of participants exposed to pesticides was overrepresented among those who had reported farming and well water consumption. However, no statistically significant association was observed between farming or well water and PD in the cohort. Several pesticides have been reported to be linked to PD, including paraquat, glyphosate, and pyrethroids [52]. Paraquat was banned in Sweden in 1983 while glyphosates and pyrethroids are still used [53]. However, further actions need to be taken to investigate which chemicals stand behind the observed association.

Having a relative with a PD diagnosis doubled the risk of having PD in the cohort. Among the 20% of patients that reported a positive family history, 8.9% had a first-degree relative with PD. Other studies report similar numbers with 15-25% of PD patients having a positive family history and 10-25% having a first-degree relative with PD [54]. It has been suggested that the risk of PD increases shortly after a traumatic brain injury (TBI) and that a history of concussions results in a higher risk of PD [55, 56]. We also observed a statistically significant association between a history of head trauma and PD, but whether the association is causative or a consequence of prodromal PD remains to be determined. Among participants reporting head trauma, no association was observed between PD and loss of consciousness.

The inverse association between smoking and PD is well-known and has been reported in numerous studies [17]. This was initially replicated in the cohort, but no longer statistically significant after adjustment for confounders, which could be due to the common co-occurrence of smoking and the use of snus. Interestingly though, a strong inverse association between the Swedish moist tobacco snus and PD was observed also after adjustment. The amount of nicotine reaching the blood when using snus is equivalent of that of cigarette smoking, and the observation reduces the number of candidate compounds underlying the inverse association between smoking and PD [57, 58]. It has previously been reported that non-smoking, snus-using Swedish men had 60% lower risk of PD compared to men who had never used snus [58]. Snus is more frequently used than smoking tobacco among men in Sweden, where 18% use snus and 7% smoke on a daily basis. Among Swedish women, 5% use snus and 7% smoke on a daily basis [59]. We investigated the total effect of smoking and snus on PD risk and found a statistically significant inverse association to PD also for the combined variable “Tobacco”. Our findings support previous reports and suggest either that components in tobacco leaves influence biological processes underlying PD, or that there is a reverse causation between tobacco use and PD. Nicotine has been suggested to have neuroprotective properties but it also stimulates dopamine release from nigrostriatal dopaminergic terminals. Hence, the lower use of tobacco products could be a consequence of a reduced nicotine-evoked dopamine release among prodromal PD patients [60]. Caffeine has also been reported to have neuroprotective effects in PD animal models, and several epidemiological studies have reported a relationship between increased coffee consumption and decreased risk of developing PD [10]. We could also observe a trend of lower OR for PD with higher coffee consumption in all investigated age groups in this cohort.

The comorbidities hypertension, hyperlipidemia, and osteoarthritis were inversely associated with PD in the cohort. Autonomic dysfunction can occur in PD, resulting in abnormalities in blood pressure, and orthostatic hypotension is common [61]. In concordance with our results, meta-analysis of PD risk factors has identified an inverse association between hypertension and PD risk [16]. However, this was only observed in case-control studies, indicating that the association might be a consequence of the disease. For hyperlipidemia, there are conflicting reports on the association with PD but a meta-analysis supports an inverse association [62]. It is possible that the use of lipid-lowering drugs (statins) contributes to the observed conflicting associations [63]. The study participants were asked whether they had been diagnosed with hyperlipidemia and, hence, it was likely that they used pharmacological treatments. Statins are the most commonly prescribed lipid-lowering drugs in Sweden, and we cannot rule out that the observed inverse association between PD and hyperlipidemia is a consequence of statin use. Moreover, the PD patients were less likely to have a diagnosis of osteoarthritis (21% patients vs 29% controls). Contrary to our observation, arthritis (no distinction was made between rheumatoid arthritis and osteoarthritis) has been reported to be the most prevalent comorbidity in PD, with almost 47% reported having the condition [64], indicating that our observation could be a consequence of underdiagnosis of osteoarthritis among the PD patients [65]. Furthermore, associations between PD and the comorbidities depression, back pain, and bowel problems were observed. Use of ibuprofen has been reported to be inversely associated with PD in a dose-dependent manner [66]. We observed a reduced PD risk for the use of the NSAID ibuprofen <2 times/week but not for ≥2 times/week and a recently published large study do not support any evidence of a decreased PD incidence among NSAID users [67].

Here, we describe to our knowledge the first GWAS of PD composed solely of PD patients from Sweden, specifically the southernmost region of Sweden. Although the relatively small sample size is a limitation in this study, issues with population stratification are expected to be lower due to region-specific study recruitment and, potentially, more homogenous ancestry. The observed near genome-wide significance associated variants at the *SNCA* locus indicate that this cohort is a well-designed case-control cohort of PD, and one of the variants was reported among the 90 risk loci in the largest GWAS meta-analysis to date [12]. Another 23 of the 90 risk loci were replicated at a nominal uncorrected p-value <0.05. Noticeable, this study had insufficient statistical power to detect variants with low MAF or small effect size, and the majority of 90 risk loci had an OR of 0.8-1.2 [12]. Insufficient statistical power is likely also the reason for the lack of an identification of any loci associated with AAD. Two genome-wide significant association signals have previously been reported to be linked to PD AAO, one at the *SNCA* locus and one at *TMEM175* [47].

Interestingly, the GWA analysis identified a novel variant, rs12771445, associated with PD at a genome-wide significance level. This variant has not been reported in the PD literature and, hence, may be specific for the Swedish population. Although, it could be that this common variant is a tagging a population-specific rare variant as the Swedish population appear to contain a substantial number of genetic variants that are not represented in other European populations [26]. However, the significant variant was imputed and we remain cautious in drawing any conclusions. As expected, we further identified one haplotype in *PLPP4* significantly associated with PD. The haplotype was more common among controls, indicating a decreased risk of PD among carriers. Furthermore, we investigated the joint effect of rare variants (MAF<5%) in *PLPP4* on PD. Due to the low coverage of coding variants, we adapted the non-burden SKAT and observed an association with PD risk for the genotyped variants but not for the imputed variants. Replication studies in larger Swedish cohorts are needed in order to validate our findings of a potential association between *PLPP4* and PD in the Swedish population.

The gene *PLPP4,* encodes for a phospholipid phosphatase that catalyzes the dephosphorylation of bioactive lipid mediators such as phosphatidic acid (PA) [68]. Another regulator of PA content, the diacylglycerol kinase, *DGKQ,* has previously been reported as a potential PD risk factor [69, 70]. The role of lipids in PD is receiving greater attention and the question whether if PD is a lipidopathy rather than a proteinopathy has been lifted [71]. Several reasons stand behind this hypothesis, including the proposal that a-synuclein is a lipid-binding protein that physiologically interacts with phospholipids and fatty acids (FAs) [71].

Although only 23 out of 90 previously reported risk variants were replicated in this Swedish cohort, the cumulative GRS based on the 90 variants showed an almost five-fold higher risk for PD in the highest quartile compared to the lowest quartile. Combining the GRS with information on age, sex, family history of PD, and the University of Pennsylvania Smell Identification Test (UPSIT) score (olfactory function) has been reported to further increase the sensitivity and specificity of the PD status model [46]. Having access to all factors but the UPSIT score, we added information on age, sex, family history of PD and PC1-5 to our model. GRS alone gave an area under the roc curve (AUC) of 0.67 while the adjusted model gave an AUC of 0.68. The non-observed difference for the two models could be due to the large proportion of the variance explained by the UPSIT score in the reported prediction model (63.1%) compared to the substantially lower values for GRS (13.6%), family history (11.4%), gender (6.0%) and age (5.9%) [46]. The lack of association to PD in the Swedish cohort for 67/90 risk variants could also have an impact on the fit of the model applied in our cohort. Although the model is functional enough to compare risk for disease status at a population level, additional information would be needed for disease prediction. Interestingly, we observed an association between the GRS and AAD despite a small sample size, with one SD increase in the Z-standardized GRS being associated with approximately one-year earlier AAD. This result is in concordance with previous results in other, larger, PD cohorts [22, 47].

Among the 90 risk variants, the *LRRK2* G2019S (rs34637584) had the largest effect estimate (2.43). This variant was genotyped in our dataset but excluded during the pre-imputation QC due to low MAF (0.2%). Previous studies of PD cohorts in Sweden have shown that the prevalence of *LRRK2* G2019S carriers is low in Sweden, 0.54% of screened 2206 PD patients were carriers [72]. It has been estimated that the G2019S mutation worldwide accounts for 4% of familial and 1% of idiopathic PD cases [73]. In our study, the fraction of carriers among the patients was 0.75%, further confirming a low prevalence of *LRRK2* G2019S carriers among Swedish PD patients.

In conclusion, this work represents a comprehensive description of a new PD case-control cohort from southern Sweden in which we nominate a novel GWA variant in the *PLPP4* locus. However, subsequent studies are needed in order to validate whether *PLPP4* is associated with PD within the Swedish population. This study contributes to the understanding of environmental and genetic risk factors in PD in the Swedish population, and the combination of epidemiological, clinical, and genetic data in the cohort makes it suitable for future studies of PD etiology.

## Data Availability

Code for all analyses and GWAS summary statistics will be available online. MultiPark's biobank sample collection (MPBC) is a resource at the Medical Faculty at Lund University which researchers can apply to for use for research within Parkinson's disease

https://github.com/KajBro/MPBC

https://drive.google.com/file/d/1kQJTIZAk1JNw-Dlu6ZHS95I2qw78Vuu_/view?usp=sharing

https://www.multipark.lu.se/infrastructures/biobank-platform

## Acknowledgments

This research was supported by MultiPark – a Strategic Research Area at Lund University and by grants from the Swedish Research Council (VR), Åke Wiberg’s foundation, Parkinsonfonden, Åhlén foundation, Lindhés advokatbyrå, and Sigurd & Elsa Golje’s memorial foundation (KB/MS). This work was also supported in part by the Intramural Research Program of the National Institute on Aging (NIA) (SBC/CB), part of the National Institutes of Health, Department of Health and Human Services. We thank the individuals who have contributed and donated blood samples to MPBC. We would also like to thank the research nurses and statistician Helene Jacobsson at MultiPark for conducting data collections, and previous students of the Translational Neurogenetics Unit who have helped with questionnaire registration to REDCap. We would like to thank Megg Garcia and Alexander Svanbergsson for technical help and feedback, and the International Parkinson’s Disease Genomic Consortium (IPDGC) and its trainee network for additional help during the project, in particularly Lynne Krohn and Manuela Tan. We would also like to acknowledge the Global Parkinson’s Genetics Program (GP2) learning platform for training courses and code in Parkinson’s disease genetics. We would also like to acknowledge Ashfaq Ali at the National Bioinformatics Infrastructure Sweden at SciLifeLab for bioinformatics advice. The Region Skåne Biobank Facility (BD47) has performed the biobank services in the project.

AP receives research support from the Swedish government (ALF), Region Skåne, Skåne University Hospital, Bundy Academy, Hans Gabriel and Alice Trolle Wachtmeister Stiftelse för Medicinsk Forskning, The Swedish Parkinson Association, The Swedish Parkinson Academy, SCA network, all in Sweden, and receives reimbursement from Elsevier Ltd. PO has received research support from The Swedish National Government and County Councils, Skåne University Hospital Foundations and Donations, Region Skåne, Parkinsonfonden, Swedish Parkinson Academy and Åhlens Foundation.

