## Supplementary material for "Insights on Genetic and Environmental Factors in Parkinson’s Disease from a regional Swedish Case-Control Cohort"

### Supplementary Data

**Table S1: Risk factors for PD in Sweden displayed as OR and 95% CI for both non-adjusted and adjusted complete-case logistic regression analyses**

| Risk factor | Control (n) | Patient (n) | Non-adjusted | | Adjusted | |
|---|---|---|---|---|---|---|
| | | | OR | 95% CI | OR | 95% CI |
| **Coffee age <41 years** | 881 | 878 | | | | |
| Nothing | 55 | 81 | 1.00 | Referent | 1.00 | Referent |
| 1-2 cups/day | 323 | 348 | 0.73 | 0.50 - 1.06 | 0.82 | 0.56 - 1.20 |
| 3-5 cups/day | 422 | 397 | 0.64 | 0.44 - 0.92 | 0.71 | 0.49 - 1.04 |
| >5 cups/day | 81 | 52 | 0.44 | 0.27 - 0.71 | 0.52 | 0.31 - 0.86 |
| **Coffee age 41-64 years** | 881 | 874 | | | | |
| Nothing | 44 | 70 | 1.00 | Referent | 1.00 | Referent |
| 1-2 cups/day | 287 | 351 | 0.77 | 0.51 - 1.15 | 0.88 | 0.57 - 1.33 |
| 3-5 cups/day | 472 | 408 | 0.54 | 0.36 - 0.81 | 0.62 | 0.41 - 0.94 |
| >5 cups/day | 78 | 45 | 0.36 | 0.21 - 0.61 | 0.43 | 0.25 - 0.74 |
| **Coffee age >64 years** | 776 | 717 | | | | |
| Nothing | 44 | 72 | 1.00 | Referent | 1.00 | Referent |
| 1-2 cups/day | 361 | 385 | 0.65 | 0.43 - 0.97 | 0.74 | 0.49 - 1.11 |
| 3-5 cups/day | 331 | 240 | 0.44 | 0.29 - 0.67 | 0.49 | 0.32 - 0.74 |
| >5 cups/day | 40 | 20 | 0.31 | 0.16 - 0.58 | 0.36 | 0.18 - 0.70 |
| **Snus** | 828 | 820 | | | | |
| Ever vs. never | 127/701 | 74/746 | 0.55 | 0.40 - 0.74 | 0.53 | 0.38 - 0.73 |
| **Tobacco** | 838 | 835 | | | | |
| Ever vs. never | 508/330 | 424/411 | 0.67 | 0.55 - 0.81 | 0.72 | 0.59 - 0.88 |
| **Smoking** | 828 | 820 | | | | |
| Current vs. never | 51/342 | 40/415 | 0.65 | 0.42 - 1.00 | 0.76 | 0.47 - 1.21 |
| Ever vs. never | 486/342 | 405/415 | 0.69 | 0.57 - 0.83 | 0.82 | 0.67 - 1.01 |
| Past vs. never | 435/342 | 365/415 | 0.69 | 0.57 - 0.84 | 0.83 | 0.67 - 1.02 |
| Pack-years (Ever-smokers) | 433 | 335 | 0.99 | 0.98 - 1.00 | 0.99 | 0.98 - 1.00 |
| **Well-water** | 847 | 738 | | | | |
| Ever vs. never | 370/477 | 334/404 | 0.94 | 0.78 - 1.13 | 1.02 | 0.83 - 1.26 |
| **BMI age 20** | 861 | 841 | 1.05 | 1.02 - 1.09 | 1.05 | 1.01 - 1.09 |
| **BMI highest** | 875 | 864 | 1.01 | 0.99 - 1.03 | 1.01 | 0.99 - 1.03 |
| **Farming** | 847 | 738 | | | | |
| Ever vs. never | 68/779 | 85/653 | 1.30 | 0.94 - 1.81 | 1.09 | 0.74 - 1.61 |
| **Head trauma** | 925 | 918 | | | | |
| Ever vs. never | 296/629 | 351/567 | 1.32 | 1.09 - 1.59 | 1.30 | 1.08 - 1.58 |
| Loss of consciousness | 133/157 | 134/209 | 0.76 | 0.55 - 1.04 | 0.76 | 0.55 - 1.04 |
| **PD family history** | 850 | 743 | | | | |
| 1st degree | 59/791 | 66/677 | 1.31 | 0.91 - 1.89 | 1.31 | 0.91 - 1.90 |
| Any relative | 93/757 | 148/595 | 2.02 | 1.53 - 2.69 | 2.00 | 1.51 - 2.67 |
| **Pesticides** | 847 | 738 | | | | |
| Ever vs. never | 31/816 | 61/677 | 2.37 | 1.53 - 3.74 | 2.26 | 1.39 - 3.72 |

**Table S2: Associations between various variables and PD.** Analyzed variables were obtained from questions regarding exposure/use within the past year prior to study inclusion. Associations are indicated in OR and 95% CI (adjusted for sex and age at inclusion).

| Variable | Control % (N) | Patient % (N) | OR | 95% CI |
|---|---|---|---|---|
| **Alcohol** | | | | |
| Nothing | 9.1 (84) | 16.1 (149) | 1.00 | Referent |
| Low | 48.2 (447) | 52.5 (481) | 0.58 | 0.43 - 0.78 |
| Moderate | 29.1 (270) | 25.1 (230) | 0.45 | 0.33 - 0.62 |
| High | 13.6 (126) | 6.2 (57) | 0.23 | 0.15 - 0.34 |
| **Red Wine** | | | | |
| Nothing | 11.8 (99) | 12.9 (99) | 1.00 | Referent |
| Low | 59.0 (497) | 63.6 (489) | 0.99 | 0.73 - 1.35 |
| Moderate | 22.0 (185) | 21.0 (161) | 0.88 | 0.62 - 1.25 |
| High | 7.2 (61) | 2.6 (20) | 0.32 | 0.18 - 0.57 |
| **BMI** | | | | |
| Inclusion | 100.0 (921) | 100.0 (914) | 0.99 | 0.96 - 1.00 |
| **Physical activity** | | | | |
| Sedentary | 7.9 (73) | 19.5 (179) | 1.00 | Referent |
| Moderate | 52.3 (484) | 51.9 (469) | 0.38 | 0.28 - 0.51 |
| Moderate but regular | 24.0 (222) | 18.2 (167) | 0.28 | 0.20 - 0.39 |
| Regular | 15.8 (146) | 11.4 (105) | 0.26 | 0.18 - 0.38 |
| **Physical activity - Hours/week** | | | | |
| Nothing | 3.2 (29) | 9.8 (84) | 1.00 | Referent |
| < 1 hour/week | 10.3 (93) | 15.0 (129) | 0.44 | 0.26 - 0.72 |
| 1-3 hours/week | 27.4 (248) | 28.8 (248) | 0.31 | 0.20 - 0.49 |
| >3 - <5 hours/week | 24.5 (222) | 21.6 (186) | 0.26 | 0.16 - 0.42 |
| ≥5 hours/week | 34.6 (313) | 24.9 (214) | 0.21 | 0.13 - 0.33 |
| **Comorbidities** | | | | |
| Hyperlipidemia | 24.6 (230/705) | 13.9 (129/800) | 0.51 | 0.40 - 0.64 |
| Hypertension | 43.7 (409/526) | 29.9 (278/651) | 0.57 | 0.47 - 0.69 |
| Osteoarthritis | 29.4 (275/660) | 21.0 (195/734) | 0.66 | 0.53 - 0.82 |
| Migraine | 7.1 (66/869) | 5.9 (55/874) | 0.81 | 0.55 - 1.17 |
| Back pain | 8.1 (76/859) | 11.8 (110/819) | 1.56 | 1.15 - 2.13 |
| Depression | 7.5 (70/865) | 13.3 (124/805) | 1.89 | 1.39 - 2.59 |
| Bowel problems | 3.5 (33/902) | 11.9 (111/818) | 3.93 | 2.66 - 5.97 |
| **Ibuprofen** | | | | |
| Never | 69.6 (514) | 78.5 (693) | 1.00 | Referent |
| < 2 times/week | 24.8 (183) | 138 (15.6) | 0.53 | 0.41 - 0.68 |
| ≥ 2 times/week | 5.7 (42) | 5.9 (52) | 0.89 | 0.58 - 1.37 |

#### Table S3: Frequency of self-reported symptoms in MPBC.

| Symptoms | Control % (N yes/no) | Patient % (N yes/no) | P-value |
|---|---|---|---|
| **Motor** | | | |
| Muscle stiffness | 10.0 (89/797) | 72.7 (647/243) | 1.23E-157 |
| Slowness of movement | 1.9 (17/863) | 72.4 (643/245) | 1.96E-205 |
| Balance problems | 11.9 (106/784) | 65.4 (581/308) | 4.53E-118 |
| Tremor | 6.0 (53/829) | 64.8 (577/313) | 6.07E-147 |
| **Non-motor** | | | |
| Nocturia | 58.4 (540/385) | 71.6 (659/261) | 3.26E-09 |
| Leg swelling | 16.5 (151/765) | 70.4 (273/648) | 3.22E-11 |
| Urgent urination | 23.5 (215/701) | 51.9 (474/440) | 9.11E-36 |
| Feeling dizzy/weak when standing up from sitting or lying down | 21.9 (200/715) | 50.2 (460/457) | 3.11E-36 |
| Reduced ability to taste or smell | 6.9 (63/852) | 45.0 (414/506) | 6.85E-77 |
| Feeling down | 17.5 (159/750) | 44.6 (410/510) | 1.32E-35 |
| Slow thinking | 12.8 (116/792) | 43.5 (396/515) | 1.17E-47 |
| Memory issues | 19.7 (178/727) | 42.3 (388/529) | 2.69E-25 |
| Unpleasant/uncomfortable sensations in the legs in the evening or when resting | 19.1 (175/739) | 41.7 (384/537) | 1.60E-25 |
| Sexual dysfunction | 28.1 (248/636) | 41.0 (350/503) | 1.70E-08 |
| Talking/moving during sleep as if “acting out” a dream | 8.1 (73/826) | 39.7 (365/555) | 2.02E-55 |
| Insomnia | 31.2 (285/628) | 39.5 (365/558) | 2.31E-04 |
| Increased or decreased libido | 28.7 (260/646) | 36.0 (319/567) | 1.13E-03 |
| Drooling | 3.3 (30/885) | 34.6 (319/603) | 3.49E-65 |
| Concentration difficulties | 6.2 (57/860) | 34.4 (316/603) | 1.77E-50 |
| Vivid dreams or nightmares | 8.8 (81/835) | 33.4 (307/612) | 1.16E-37 |
| Constipation | 5.6 (51/865) | 33.3 (305/612) | 2.22E-50 |
| Falling | 7.0 (64/850) | 30.3 (278/638) | 3.10E-37 |
| Feeling of incomplete evacuation of stools | 11.2 (102/812) | 28.6 (261/653) | 1.98E-20 |
| Idiopathic pain | 7.8 (71/841) | 26.9 (245/666) | 8.70E-27 |
| Feeling anxious/worried/scared or panicky | 6.7 (61/846) | 26.0 (239/681) | 2.36E-28 |
| Difficulties swallowing | 5.9 (54/862) | 24.6 (226/691) | 1.37E-28 |
| Lost interest | 3.1 (28/889) | 21.6 (198/720) | 3.70E-33 |
| Excessive sweating | 8.4 (77/838) | 18.8 (173/746) | 1.29E-10 |
| Visual-spatial problem | 2.7 (25/887) | 17.4 (159/755) | 5.49E-25 |
| Diplopia | 2.1 (19/888) | 17.1 (157/760) | 3.93E-27 |
| Hallucinations | 1.1 (10/904) | 16.8 (155/765) | 1.17E-31 |
| Hypersomnia | 2.4 (22/897) | 13.0 (120/806) | 3.59E-17 |
| Unexplained weight loss | 1.4 (13/900) | 11.2 (103/815) | 1.77E-17 |
| Fecal incontinence | 3.4 (31/884) | 7.3 (66/834) | 2.81E-04 |
| Nausea | 1.8 (16/898) | 7.0 (64/854) | 8.63E-08 |
| Delusions | 0.5 (5/909) | 6.0 (55/860) | 1.29E-10 |

**Table S4: Top variants associated with PD in MPBC from GWA analysis.** Table showing the results from GWA analysis in MPBC along with information from summary statistics on the variants retrieved from the latest GWAS meta-analysis of individuals with European ancestry.

| Nearest gene(s) | SNP | CHR | POS | Effect allele | Alt. allele | EAF | MAF | Genotyped | Rsq | Beta | OR | SE | P | EAF[a] | MAF[a] | Beta[a] | OR | SE[a] | P[a] |
|---|---|---|---|---|---|---|---|---|---|---|---|---|---|---|---|---|---|---|---|
| **PLPP4** | **rs12771445** | **10** | **122318147** | **T** | **C** | **0.314** | **0.314** | **Imputed** | **0.986** | **-0.411** | **0.663** | **0.072** | **1.30E-08** | 0.324 | 0.324 | 0.015 | 1.015 | 0.021 | 4.88E-01 |
| SNCA | rs356182 | 4 | 90626111 | A | G | 0.617 | 0.383 | Genotyped | 0.997 | -0.377 | 0.686 | 0.069 | 5.64E-08 | 0.616 | 0.384 | -0.255 | 0.775 | 0.021 | 9.41E-34 |
| SNCA | rs356203 | 4 | 90666041 | T | C | 0.588 | 0.412 | Imputed | 0.977 | -0.362 | 0.697 | 0.068 | 1.27E-07 | 0.617 | 0.383 | -0.240 | 0.787 | 0.018 | 3.01E-41 |
| SNCA/ LOC105377329 | rs356220 | 4 | 90641340 | T | C | 0.407 | 0.407 | Genotyped | 1.000 | 0.358 | 1.430 | 0.068 | 1.39E-07 | 0.390 | 0.390 | 0.233 | 1.262 | 0.017 | 3.92E-41 |
| PLPP4 | rs11596921 | 10 | 122339965 | C | G | 0.322 | 0.322 | Imputed | 0.989 | -0.378 | 0.685 | 0.072 | 1.45E-07 | 0.341 | 0.341 | 0.002 | 1.002 | 0.019 | 9.33E-01 |
| SNCA/ LOC105377329 | rs356219 | 4 | 90637601 | A | G | 0.590 | 0.410 | Imputed | 0.976 | -0.355 | 0.701 | 0.069 | 2.32E-07 | 0.611 | 0.389 | -0.233 | 0.792 | 0.017 | 3.47E-41 |
| COL12A1/ LOC105377858 | rs7752646 | 6 | 75447088 | T | G | 0.873 | 0.127 | Imputed | 0.958 | -0.715 | 0.489 | 0.106 | 4.13E-07 | 0.862 | 0.139 | -0.060 | 0.942 | 0.033 | 6.62E-02 |

a = Nalls 2019 without 23andMe data [12]

SNCA rs356182 can also be found in table S5

### Table S5: Results from GWA analysis in MPBC for the 90 risk variants reported to be associated with PD in cohorts of European ancestry

| Nearest gene(s) | SNP | CHR | POS | Effect allele | Alt. allele | EAF | MAF | Genotyped | Rsq | Beta | OR | SE | P | EAF[a] | MAF[a] | Beta[a] | OR[a] | SE[a] | P[a] | EAF[b] | MAF[b] | Beta[b] | OR[b] | SE[b] | P[b] |
|---|---|---|---|---|---|---|---|---|---|---|---|---|---|---|---|---|---|---|---|---|---|---|---|---|---|
| PMVK | rs114138760 | 1 | 154898185 | C | G | 0.007 | 0.007 | Imputed | 0.914 | -0.136 | 0.873 | 0.411 | 7.40E-01 | 0.011 | 0.011 | 0.311 | 1.365 | 0.084 | 2.25E-04 | 0.011 | 0.011 | 0.281 | 1.324 | 0.048 | 4.19E-09 |
| KRTCAP2 | rs35749011 | 1 | 155135036 | A | G | 0.024 | 0.024 | Imputed | 0.997 | 0.649 | 1.914 | 0.229 | 4.52E-03 | 0.019 | 0.019 | 0.751 | 2.119 | 0.066 | 5.02E-30 | 0.017 | 0.017 | 0.607 | 1.835 | 0.034 | 1.72E-70 |
| GBAP1 | rs76763715 | 1 | 155205634 | T | C | 0.998 | 0.002 | Imputed | 0.826 | -1.832 | 0.160 | 1.026 | 7.42E-02 | 0.993 | 0.007 | -0.491 | 0.612 | 0.143 | 5.76E-04 | 0.995 | 0.005 | -0.747 | 0.474 | 0.077 | 1.59E-22 |
| FCGR2A | rs6658353 | 1 | 161469054 | C | G | 0.505 | 0.495 | Imputed | 0.986 | 0.120 | 1.127 | 0.066 | 6.92E-02 | 0.501 | 0.499 | 0.072 | 1.075 | 0.017 | 2.42E-05 | 0.501 | 0.499 | 0.065 | 1.067 | 0.009 | 6.10E-12 |
| VAMP4 | rs11578699 | 1 | 171719769 | T | C | 0.179 | 0.179 | Imputed | 0.988 | -0.034 | 0.967 | 0.087 | 7.00E-01 | 0.196 | 0.196 | -0.078 | 0.925 | 0.022 | 4.24E-04 | 0.195 | 0.195 | -0.070 | 0.932 | 0.012 | 4.47E-09 |
| NUCKS1 | rs823118 | 1 | 205723572 | C | T | 0.560 | 0.440 | Genotyped | 1.000 | 0.141 | 1.151 | 0.066 | 3.37E-02 | 0.575 | 0.425 | 0.100 | 1.105 | 0.017 | 4.94E-09 | 0.566 | 0.434 | 0.107 | 1.113 | 0.009 | 1.11E-29 |
| RAB29 | rs11557080 | 1 | 205737739 | A | G | 0.109 | 0.109 | Imputed | 0.972 | 0.194 | 1.214 | 0.108 | 7.19E-02 | 0.143 | 0.143 | 0.135 | 1.145 | 0.024 | 2.12E-08 | 0.139 | 0.139 | 0.132 | 1.141 | 0.014 | 2.50E-22 |
| ITPKB | rs4653767 | 1 | 226916078 | T | C | 0.739 | 0.261 | Imputed | 0.991 | 0.022 | 1.022 | 0.076 | 7.76E-01 | 0.716 | 0.284 | 0.073 | 1.076 | 0.019 | 8.67E-05 | 0.720 | 0.280 | 0.083 | 1.087 | 0.010 | 1.38E-15 |
| SIPA1L2 | rs10797576 | 1 | 232664611 | T | C | 0.107 | 0.107 | Genotyped | 0.999 | 0.122 | 1.130 | 0.106 | 2.48E-01 | 0.143 | 0.143 | 0.100 | 1.105 | 0.024 | 3.53E-05 | 0.140 | 0.140 | 0.111 | 1.117 | 0.013 | 6.84E-17 |
| KCNS3 | rs76116224 | 2 | 18147848 | A | T | 0.895 | 0.105 | Imputed | 0.952 | 0.154 | 1.166 | 0.109 | 1.58E-01 | 0.911 | 0.090 | 0.155 | 1.168 | 0.040 | 1.19E-04 | 0.904 | 0.096 | 0.110 | 1.116 | 0.019 | 1.27E-08 |
| KCNIP3 | rs2042477 | 2 | 96000943 | A | T | 0.241 | 0.241 | Imputed | 0.877 | 0.048 | 1.049 | 0.078 | 5.43E-01 | 0.237 | 0.237 | -0.058 | 0.944 | 0.022 | 6.89E-03 | 0.242 | 0.242 | -0.066 | 0.936 | 0.012 | 1.38E-08 |
| MAP4K4 | rs11683001 | 2 | 102396963 | A | T | 0.339 | 0.339 | Imputed | 0.987 | 0.068 | 1.070 | 0.070 | 3.30E-01 | 0.332 | 0.332 | 0.076 | 1.079 | 0.018 | 2.11E-05 | 0.337 | 0.337 | 0.071 | 1.074 | 0.010 | 8.04E-13 |
| TMEM163 | rs57891859 | 2 | 135464616 | A | G | 0.768 | 0.232 | Imputed | 0.976 | 0.097 | 1.102 | 0.079 | 2.23E-01 | 0.715 | 0.285 | 0.111 | 1.118 | 0.019 | 4.93E-09 | 0.719 | 0.281 | 0.081 | 1.084 | 0.011 | 4.55E-14 |
| STK39 | rs1474055 | 2 | 169110394 | T | C | 0.123 | 0.123 | Imputed | 0.985 | 0.140 | 1.150 | 0.101 | 1.63E-01 | 0.133 | 0.133 | 0.176 | 1.193 | 0.025 | 1.14E-12 | 0.131 | 0.131 | 0.180 | 1.197 | 0.014 | 2.54E-39 |
| SATB1 | rs73038319 | 3 | 18361759 | A | C | 0.933 | 0.067 | Imputed | 0.972 | -0.048 | 0.953 | 0.135 | 7.19E-01 | 0.961 | 0.039 | -0.195 | 0.823 | 0.045 | 1.30E-05 | 0.959 | 0.041 | -0.169 | 0.845 | 0.024 | 5.94E-13 |
| LINC00693 | rs6808178 | 3 | 28705690 | T | C | 0.390 | 0.390 | Imputed | 0.972 | 0.165 | 1.179 | 0.068 | 1.55E-02 | 0.377 | 0.377 | 0.086 | 1.090 | 0.017 | 7.20E-07 | 0.379 | 0.379 | 0.066 | 1.068 | 0.010 | 8.09E-12 |
| IP6K2 | rs12497850 | 3 | 48748989 | T | C | 0.619 | 0.381 | Imputed | 0.976 | -0.018 | 0.982 | 0.068 | 7.92E-01 | 0.647 | 0.353 | 0.049 | 1.050 | 0.018 | 5.74E-03 | 0.648 | 0.352 | 0.064 | 1.066 | 0.010 | 1.36E-10 |
| KPNA1 | rs55961674 | 3 | 122196892 | T | C | 0.134 | 0.134 | Imputed | 0.930 | 0.106 | 1.112 | 0.100 | 2.90E-01 | 0.179 | 0.179 | 0.083 | 1.087 | 0.023 | 2.49E-04 | 0.172 | 0.172 | 0.086 | 1.090 | 0.013 | 9.98E-12 |
| MED12L | rs11707416 | 3 | 151108965 | A | T | 0.375 | 0.375 | Imputed | 0.994 | -0.051 | 0.950 | 0.067 | 4.48E-01 | 0.370 | 0.370 | -0.072 | 0.931 | 0.018 | 4.53E-05 | 0.367 | 0.367 | -0.063 | 0.939 | 0.010 | 1.13E-10 |
| SPTSSB | rs1450522 | 3 | 161077630 | A | G | 0.678 | 0.322 | Imputed | 0.995 | 0.029 | 1.029 | 0.071 | 6.89E-01 | 0.673 | 0.327 | -0.047 | 0.954 | 0.018 | 8.63E-03 | 0.674 | 0.326 | -0.062 | 0.940 | 0.010 | 5.01E-10 |
| MCCC1 | rs10513789 | 3 | 182760073 | T | G | 0.783 | 0.217 | Genotyped | 1.000 | 0.156 | 1.169 | 0.081 | 5.38E-02 | 0.817 | 0.183 | 0.160 | 1.173 | 0.022 | 3.19E-13 | 0.811 | 0.189 | 0.149 | 1.161 | 0.012 | 1.22E-34 |
| GAK | rs873786 | 4 | 925376 | T | C | 0.102 | 0.102 | Imputed | 0.948 | -0.281 | 0.755 | 0.114 | 1.38E-02 | 0.100 | 0.100 | -0.135 | 0.874 | 0.030 | 8.70E-06 | 0.099 | 0.099 | -0.173 | 0.841 | 0.018 | 1.79E-21 |
| TMEM175 | rs34311866 | 4 | 951947 | T | C | 0.803 | 0.197 | Imputed | 0.986 | -0.332 | 0.717 | 0.085 | 1.02E-04 | 0.804 | 0.196 | -0.227 | 0.797 | 0.023 | 7.97E-23 | 0.807 | 0.193 | -0.213 | 0.808 | 0.012 | 9.98E-70 |
| BST1 | rs4698412 | 4 | 15737348 | A | G | 0.564 | 0.436 | Imputed | 0.980 | 0.098 | 1.103 | 0.067 | 1.44E-01 | 0.553 | 0.447 | 0.126 | 1.134 | 0.017 | 7.05E-14 | 0.553 | 0.447 | 0.104 | 1.110 | 0.009 | 2.06E-28 |
| LCORL | rs34025766 | 4 | 17968811 | A | T | 0.129 | 0.129 | Imputed | 0.997 | -0.217 | 0.805 | 0.098 | 2.66E-02 | 0.162 | 0.162 | -0.068 | 0.934 | 0.024 | 4.57E-03 | 0.159 | 0.159 | -0.084 | 0.919 | 0.013 | 2.87E-10 |
| SCARB2 | rs6825004 | 4 | 77110365 | C | G | 0.706 | 0.294 | Imputed | 0.975 | 0.116 | 1.123 | 0.073 | 1.11E-01 | 0.692 | 0.308 | 0.035 | 1.035 | 0.018 | 6.05E-02 | 0.691 | 0.309 | 0.062 | 1.064 | 0.010 | 1.17E-09 |
| FAM47E | rs4101061 | 4 | 77147969 | A | G | 0.707 | 0.293 | Imputed | 0.987 | -0.164 | 0.849 | 0.073 | 2.40E-02 | 0.715 | 0.286 | -0.096 | 0.909 | 0.019 | 2.96E-07 | 0.711 | 0.289 | -0.091 | 0.913 | 0.010 | 4.97E-19 |
| FAM47E-STBD1 | rs6854006 | 4 | 77198054 | T | C | 0.353 | 0.353 | Imputed | 0.996 | -0.187 | 0.829 | 0.069 | 6.65E-03 | 0.363 | 0.363 | -0.097 | 0.908 | 0.018 | 3.50E-08 | 0.363 | 0.363 | -0.091 | 0.913 | 0.010 | 5.82E-21 |
| SNCA | rs356182* | 4 | 90626111 | A | G | 0.617 | 0.383 | Genotyped | 0.997 | -0.377 | 0.686 | 0.069 | 5.64E-08 | 0.616 | 0.384 | -0.255 | 0.775 | 0.021 | 9.41E-34 | 0.628 | 0.372 | -0.277 | 0.758 | 0.011 | 3.89E-154 |
| SNCA | rs5019538 | 4 | 90636630 | A | G | 0.689 | 0.311 | Imputed | 0.971 | -0.206 | 0.814 | 0.071 | 3.65E-03 | 0.687 | 0.313 | -0.169 | 0.844 | 0.018 | 2.82E-20 | 0.679 | 0.321 | -0.157 | 0.855 | 0.012 | 1.13E-36 |
| CAMK2D | rs13117519 | 4 | 114369065 | T | C | 0.184 | 0.184 | Imputed | 0.958 | 0.103 | 1.108 | 0.089 | 2.46E-01 | 0.179 | 0.179 | 0.072 | 1.075 | 0.022 | 1.10E-03 | 0.174 | 0.174 | 0.088 | 1.092 | 0.012 | 9.82E-13 |
| CLCN3 | rs62333164 | 4 | 170583157 | A | G | 0.326 | 0.326 | Imputed | 0.976 | -0.072 | 0.931 | 0.070 | 3.03E-01 | 0.322 | 0.322 | -0.058 | 0.943 | 0.018 | 1.44E-03 | 0.326 | 0.326 | -0.064 | 0.938 | 0.010 | 2.00E-10 |
| ELOVL7 | rs1867598 | 5 | 60137959 | A | G | 0.901 | 0.099 | Imputed | 0.994 | -0.018 | 0.982 | 0.114 | 8.78E-01 | 0.899 | 0.101 | -0.198 | 0.821 | 0.028 | 8.74E-13 | 0.902 | 0.098 | -0.155 | 0.856 | 0.016 | 2.52E-23 |
| PAM | rs26431 | 5 | 102365794 | C | G | 0.718 | 0.282 | Imputed | 0.934 | 0.123 | 1.131 | 0.076 | 1.06E-01 | 0.701 | 0.300 | 0.063 | 1.065 | 0.018 | 5.37E-04 | 0.703 | 0.297 | 0.062 | 1.064 | 0.010 | 1.57E-09 |
| C5orf24 | rs11950533 | 5 | 134199105 | A | C | 0.122 | 0.122 | Imputed | 0.946 | -0.150 | 0.861 | 0.107 | 1.61E-01 | 0.101 | 0.101 | -0.088 | 0.915 | 0.028 | 1.81E-03 | 0.102 | 0.102 | -0.092 | 0.912 | 0.016 | 7.16E-09 |
| LOC100131289 | rs4140646 | 6 | 27738801 | A | G | 0.245 | 0.245 | Imputed | 0.995 | 0.091 | 1.095 | 0.075 | 2.23E-01 | 0.222 | 0.222 | 0.073 | 1.076 | 0.022 | 8.35E-04 | 0.208 | 0.208 | 0.083 | 1.087 | 0.012 | 5.62E-12 |
| TRIM40 | rs9261484 | 6 | 30108683 | T | C | 0.284 | 0.284 | Genotyped | 1.000 | -0.095 | 0.909 | 0.073 | 1.91E-01 | 0.240 | 0.240 | -0.046 | 0.956 | 0.021 | 3.26E-02 | 0.245 | 0.245 | -0.064 | 0.938 | 0.011 | 1.62E-08 |
| HLA-DRB5 | rs112485576 | 6 | 32578772 | A | C | 0.186 | 0.186 | Imputed | 0.993 | -0.175 | 0.839 | 0.084 | 3.78E-02 | 0.155 | 0.155 | -0.187 | 0.829 | 0.029 | 1.36E-10 | 0.163 | 0.163 | -0.168 | 0.845 | 0.015 | 6.96E-28 |
| RIMS1 | rs12528068 | 6 | 72487762 | T | C | 0.296 | 0.296 | Imputed | 0.992 | -0.025 | 0.975 | 0.073 | 7.31E-01 | 0.286 | 0.286 | 0.083 | 1.086 | 0.019 | 8.37E-06 | 0.284 | 0.284 | 0.066 | 1.068 | 0.010 | 1.63E-10 |
| FYN | rs997368 | 6 | 112243291 | A | G | 0.815 | 0.185 | Imputed | 0.976 | 0.026 | 1.026 | 0.085 | 7.62E-01 | 0.801 | 0.199 | 0.053 | 1.055 | 0.021 | 1.19E-02 | 0.805 | 0.195 | 0.071 | 1.074 | 0.012 | 1.84E-09 |
| RPS12 | rs75859381 | 6 | 133210361 | T | C | 0.954 | 0.046 | Imputed | 0.965 | -0.261 | 0.770 | 0.165 | 1.14E-01 | 0.970 | 0.030 | -0.285 | 0.752 | 0.067 | 1.95E-05 | 0.967 | 0.033 | -0.221 | 0.802 | 0.034 | 1.04E-10 |
| GPNMB | rs199351 | 7 | 23300049 | A | C | 0.607 | 0.393 | Imputed | 0.989 | 0.058 | 1.060 | 0.067 | 3.84E-01 | 0.591 | 0.409 | 0.099 | 1.104 | 0.017 | 1.28E-08 | 0.594 | 0.406 | 0.102 | 1.107 | 0.010 | 5.25E-26 |
| GS1-124K5.11 | rs76949143 | 7 | 66009851 | A | T | 0.067 | 0.067 | Imputed | 0.985 | -0.357 | 0.700 | 0.135 | 8.21E-03 | 0.058 | 0.058 | -0.118 | 0.889 | 0.052 | 2.23E-02 | 0.051 | 0.051 | -0.143 | 0.867 | 0.025 | 1.43E-08 |
| CTSB | rs1293298 | 8 | 11712443 | A | C | 0.759 | 0.241 | Imputed | 0.929 | 0.151 | 1.163 | 0.081 | 6.12E-02 | 0.749 | 0.251 | 0.089 | 1.093 | 0.022 | 3.73E-05 | 0.744 | 0.256 | 0.093 | 1.097 | 0.011 | 3.99E-16 |
| FGF20 | rs620513 | 8 | 16697593 | T | G | 0.274 | 0.274 | Imputed | 0.979 | -0.060 | 0.942 | 0.074 | 4.15E-01 | 0.268 | 0.268 | -0.115 | 0.892 | 0.019 | 2.14E-09 | 0.268 | 0.268 | -0.086 | 0.918 | 0.011 | 2.72E-15 |
| BIN3 | rs2280104 | 8 | 22525980 | T | C | 0.369 | 0.369 | Genotyped | 0.999 | 0.049 | 1.050 | 0.068 | 4.72E-01 | 0.364 | 0.364 | 0.061 | 1.063 | 0.018 | 4.53E-04 | 0.360 | 0.360 | 0.056 | 1.058 | 0.010 | 1.16E-08 |
| FAM49B | rs2086641 | 8 | 130901909 | T | C | 0.746 | 0.254 | Imputed | 0.955 | -0.009 | 0.991 | 0.077 | 9.10E-01 | 0.721 | 0.279 | -0.068 | 0.934 | 0.021 | 1.11E-03 | 0.723 | 0.277 | -0.061 | 0.941 | 0.011 | 1.81E-08 |
| SH3GL2 | rs13294100 | 9 | 17579690 | T | G | 0.356 | 0.356 | Imputed | 0.969 | -0.251 | 0.778 | 0.070 | 3.62E-04 | 0.341 | 0.341 | -0.086 | 0.918 | 0.018 | 1.20E-06 | 0.342 | 0.342 | -0.086 | 0.918 | 0.010 | 8.72E-18 |
| SH3GL2 | rs10756907 | 9 | 17727065 | A | G | 0.733 | 0.267 | Imputed | 0.957 | -0.224 | 0.799 | 0.075 | 2.86E-03 | 0.760 | 0.240 | -0.100 | 0.905 | 0.020 | 3.10E-07 | 0.767 | 0.233 | -0.093 | 0.911 | 0.011 | 5.06E-17 |
| UBAP2 | rs6476434 | 9 | 34046391 | T | C | 0.741 | 0.259 | Imputed | 0.956 | -0.022 | 0.978 | 0.075 | 7.73E-01 | 0.728 | 0.273 | -0.043 | 0.958 | 0.019 | 2.52E-02 | 0.734 | 0.266 | -0.062 | 0.940 | 0.011 | 6.58E-09 |
| ITGA8 | rs896435 | 10 | 15557406 | T | C | 0.681 | 0.319 | Imputed | 0.968 | -0.092 | 0.912 | 0.072 | 2.03E-01 | 0.690 | 0.310 | 0.055 | 1.056 | 0.018 | 2.61E-03 | 0.689 | 0.311 | 0.074 | 1.077 | 0.010 | 3.41E-13 |
| GBF1 | rs10748818 | 10 | 104015279 | A | G | 0.840 | 0.160 | Imputed | 0.986 | -0.077 | 0.926 | 0.089 | 3.87E-01 | 0.852 | 0.148 | -0.050 | 0.952 | 0.024 | 3.59E-02 | 0.851 | 0.149 | -0.079 | 0.924 | 0.013 | 1.05E-09 |
| BAG3 | rs72840788 | 10 | 121415685 | A | G | 0.248 | 0.248 | Imputed | 0.981 | -0.061 | 0.941 | 0.077 | 4.27E-01 | 0.217 | 0.217 | 0.091 | 1.095 | 0.021 | 1.00E-05 | 0.216 | 0.216 | 0.076 | 1.079 | 0.011 | 1.57E-11 |
| INPP5F | rs117896735 | 10 | 121536327 | A | G | 0.016 | 0.016 | Imputed | 0.946 | 0.811 | 2.250 | 0.293 | 5.73E-03 | 0.018 | 0.018 | 0.419 | 1.521 | 0.067 | 2.83E-10 | 0.017 | 0.017 | 0.435 | 1.545 | 0.039 | 2.36E-28 |
| RNF141 | rs7938782 | 11 | 10558777 | A | G | 0.899 | 0.101 | Imputed | 0.981 | 0.370 | 1.448 | 0.114 | 1.14E-03 | 0.873 | 0.127 | 0.077 | 1.080 | 0.026 | 2.94E-03 | 0.878 | 0.122 | 0.087 | 1.091 | 0.015 | 2.12E-09 |
| DLG2 | rs12283611 | 11 | 83487277 | A | C | 0.391 | 0.391 | Imputed | 0.974 | -0.031 | 0.969 | 0.068 | 6.46E-01 | 0.417 | 0.417 | -0.050 | 0.951 | 0.017 | 3.23E-03 | 0.415 | 0.415 | -0.065 | 0.937 | 0.010 | 2.61E-10 |
| IGSF9B | rs3802920 | 11 | 133787001 | T | G | 0.164 | 0.164 | Imputed | 0.992 | 0.107 | 1.113 | 0.090 | 2.35E-01 | 0.212 | 0.212 | 0.112 | 1.119 | 0.021 | 1.65E-07 | 0.205 | 0.205 | 0.107 | 1.113 | 0.012 | 6.26E-20 |
| LRRK2 | rs76904798 | 12 | 40614434 | T | C | 0.126 | 0.126 | Genotyped | 1.000 | 0.265 | 1.303 | 0.101 | 8.67E-03 | 0.149 | 0.149 | 0.135 | 1.145 | 0.024 | 9.22E-09 | 0.144 | 0.144 | 0.144 | 1.155 | 0.013 | 1.52E-28 |
| LRRK2 | rs34637584 | 12 | 40734202 | A | G | 0.001 | 0.001 | Imputed | 0.822 | 1.661 | 5.265 | 1.255 | 1.86E-01 | 0.005 | 0.005 | 2.124 | 8.365 | 0.300 | 1.33E-12 | 0.002 | 0.002 | 2.429 | 11.348 | 0.094 | 3.61E-148 |
| *LRRK2* | *rs34637584* | *12* | *40734202* | *A* | *G* | *0.002* | *0.002* | *Genotyped* | - | - | - | - | - | *0.005* | *0.005* | *2.124* | *8.365* | *0.300* | *1.33E-12* | *0.002* | *0.002* | *2.429* | *11.348* | *0.094* | *3.61E-148* |
| SCAF11 | rs7134559 | 12 | 46419086 | T | C | 0.353 | 0.353 | Imputed | 0.985 | -0.030 | 0.970 | 0.071 | 6.72E-01 | 0.402 | 0.402 | -0.074 | 0.928 | 0.017 | 1.51E-05 | 0.404 | 0.404 | -0.054 | 0.947 | 0.010 | 3.96E-08 |
| HIP1R | rs10847864 | 12 | 123326598 | T | G | 0.342 | 0.342 | Imputed | 0.913 | 0.158 | 1.171 | 0.073 | 3.09E-02 | 0.363 | 0.363 | 0.127 | 1.136 | 0.018 | 9.81E-13 | 0.364 | 0.364 | 0.148 | 1.160 | 0.012 | 1.47E-37 |
| FBRSL1 | rs11610045 | 12 | 133063768 | A | G | 0.554 | 0.446 | Genotyped | 0.999 | 0.141 | 1.151 | 0.067 | 3.61E-02 | 0.482 | 0.482 | 0.057 | 1.058 | 0.017 | 7.40E-04 | 0.490 | 0.490 | 0.060 | 1.062 | 0.009 | 1.77E-10 |
| CAB39L | rs9568188 | 13 | 49927732 | T | C | 0.677 | 0.323 | Imputed | 0.993 | 0.040 | 1.041 | 0.070 | 5.72E-01 | 0.744 | 0.256 | 0.024 | 1.024 | 0.019 | 2.27E-01 | 0.740 | 0.260 | 0.062 | 1.064 | 0.011 | 1.15E-08 |
| MBNL2 | rs4771268 | 13 | 97865021 | T | C | 0.218 | 0.218 | Imputed | 0.940 | -0.021 | 0.979 | 0.083 | 7.95E-01 | 0.232 | 0.232 | 0.072 | 1.075 | 0.020 | 2.74E-04 | 0.230 | 0.230 | 0.068 | 1.070 | 0.011 | 1.45E-09 |
| MIPOL1 | rs12147950 | 14 | 37989270 | T | C | 0.417 | 0.417 | Imputed | 0.950 | -0.005 | 0.995 | 0.068 | 9.46E-01 | 0.436 | 0.436 | -0.079 | 0.924 | 0.018 | 9.46E-06 | 0.438 | 0.438 | -0.053 | 0.948 | 0.010 | 3.54E-08 |
| GCH1 | rs11158026 | 14 | 55348869 | T | C | 0.326 | 0.326 | Genotyped | 0.999 | -0.138 | 0.871 | 0.070 | 4.94E-02 | 0.316 | 0.316 | -0.066 | 0.936 | 0.018 | 2.59E-04 | 0.325 | 0.325 | -0.084 | 0.919 | 0.010 | 1.66E-16 |
| RPS6KL1 | rs3742785 | 14 | 75373034 | A | C | 0.769 | 0.231 | Imputed | 0.988 | 0.006 | 1.006 | 0.078 | 9.34E-01 | 0.786 | 0.214 | 0.083 | 1.086 | 0.023 | 2.60E-04 | 0.787 | 0.213 | 0.071 | 1.074 | 0.012 | 1.92E-09 |
| GALC | rs979812 | 14 | 88464264 | T | G | 0.430 | 0.430 | Imputed | 0.981 | 0.040 | 1.041 | 0.067 | 5.55E-01 | 0.445 | 0.445 | 0.036 | 1.036 | 0.017 | 3.44E-02 | 0.442 | 0.442 | 0.061 | 1.063 | 0.009 | 6.19E-11 |
| VPS13C | rs2251086 | 15 | 61997385 | T | C | 0.144 | 0.144 | Imputed | 0.961 | -0.103 | 0.902 | 0.094 | 2.73E-01 | 0.135 | 0.135 | -0.128 | 0.880 | 0.025 | 2.53E-07 | 0.142 | 0.142 | -0.119 | 0.888 | 0.014 | 6.08E-18 |
| SYT17 | rs6497339 | 16 | 19277493 | A | T | 0.504 | 0.497 | Imputed | 0.923 | 0.013 | 1.013 | 0.068 | 8.54E-01 | 0.461 | 0.461 | 0.045 | 1.046 | 0.017 | 8.61E-03 | 0.454 | 0.454 | 0.063 | 1.065 | 0.010 | 2.76E-11 |
| CD19 | rs2904880 | 16 | 28944396 | C | G | 0.333 | 0.333 | Imputed | 0.921 | 0.056 | 1.058 | 0.071 | 4.32E-01 | 0.305 | 0.305 | -0.066 | 0.936 | 0.019 | 4.49E-04 | 0.309 | 0.309 | -0.065 | 0.937 | 0.011 | 7.87E-10 |
| SETD1A | rs11150601 | 16 | 30977799 | A | G | 0.642 | 0.358 | Imputed | 0.960 | 0.062 | 1.064 | 0.069 | 3.67E-01 | 0.653 | 0.347 | 0.112 | 1.119 | 0.018 | 9.02E-10 | 0.644 | 0.356 | 0.091 | 1.095 | 0.010 | 5.12E-20 |
| NOD2 | rs6500328 | 16 | 50736656 | A | G | 0.593 | 0.407 | Imputed | 0.990 | 0.012 | 1.012 | 0.067 | 8.57E-01 | 0.602 | 0.398 | 0.070 | 1.073 | 0.018 | 1.35E-04 | 0.599 | 0.401 | 0.059 | 1.061 | 0.010 | 1.82E-09 |
| CASC16 | rs3104783 | 16 | 52636242 | A | C | 0.376 | 0.376 | Imputed | 0.974 | 0.046 | 1.047 | 0.070 | 5.13E-01 | 0.437 | 0.437 | 0.081 | 1.084 | 0.017 | 1.94E-06 | 0.434 | 0.434 | 0.067 | 1.069 | 0.009 | 1.29E-12 |
| CHD9 | rs10221156 | 16 | 52969426 | A | G | 0.091 | 0.091 | Imputed | 0.986 | -0.222 | 0.801 | 0.116 | 5.55E-02 | 0.090 | 0.090 | -0.162 | 0.851 | 0.040 | 5.75E-05 | 0.093 | 0.093 | -0.116 | 0.890 | 0.018 | 1.08E-10 |
| CHRNB1 | rs12600861 | 17 | 7355621 | A | C | 0.619 | 0.381 | Imputed | 0.974 | -0.046 | 0.955 | 0.069 | 5.09E-01 | 0.645 | 0.355 | -0.062 | 0.940 | 0.018 | 5.10E-04 | 0.648 | 0.352 | -0.057 | 0.945 | 0.010 | 1.01E-08 |
| RETREG3 | rs12951632 | 17 | 40741013 | T | C | 0.724 | 0.276 | Imputed | 0.983 | 0.186 | 1.204 | 0.076 | 1.41E-02 | 0.733 | 0.267 | 0.072 | 1.075 | 0.019 | 1.35E-04 | 0.735 | 0.265 | 0.064 | 1.066 | 0.011 | 1.40E-09 |
| UBTF | rs2269906 | 17 | 42294337 | A | C | 0.696 | 0.304 | Imputed | 0.987 | 0.104 | 1.110 | 0.072 | 1.52E-01 | 0.660 | 0.340 | 0.061 | 1.063 | 0.020 | 1.90E-03 | 0.653 | 0.347 | 0.063 | 1.065 | 0.010 | 6.24E-10 |
| FAM171A2 | rs850738 | 17 | 42434630 | A | G | 0.576 | 0.424 | Imputed | 0.956 | -0.117 | 0.890 | 0.069 | 8.96E-02 | 0.604 | 0.396 | -0.082 | 0.921 | 0.022 | 1.89E-04 | 0.606 | 0.394 | -0.071 | 0.931 | 0.011 | 1.29E-11 |
| CRHR1 | rs62053943 | 17 | 43744203 | T | C | 0.110 | 0.110 | Imputed | 0.881 | -0.156 | 0.856 | 0.112 | 1.63E-01 | 0.145 | 0.145 | -0.229 | 0.795 | 0.031 | 5.86E-14 | 0.155 | 0.155 | -0.270 | 0.763 | 0.016 | 3.58E-68 |
| CRHR1 | rs117615688 | 17 | 43798308 | A | G | 0.040 | 0.040 | Imputed | 0.729 | -0.410 | 0.664 | 0.194 | 3.46E-02 | 0.072 | 0.072 | -0.164 | 0.849 | 0.042 | 1.07E-04 | 0.067 | 0.067 | -0.232 | 0.793 | 0.029 | 6.71E-16 |
| WNT3 | rs11658976 | 17 | 44866805 | A | G | 0.584 | 0.416 | Genotyped | 0.993 | -0.087 | 0.917 | 0.067 | 1.96E-01 | 0.603 | 0.397 | -0.061 | 0.941 | 0.023 | 7.81E-03 | 0.580 | 0.420 | -0.062 | 0.940 | 0.011 | 3.52E-08 |
| BRIP1 | rs61169879 | 17 | 59917366 | T | C | 0.173 | 0.173 | Imputed | 0.973 | -0.038 | 0.963 | 0.088 | 6.64E-01 | 0.167 | 0.167 | 0.067 | 1.069 | 0.030 | 2.45E-02 | 0.164 | 0.164 | 0.082 | 1.085 | 0.013 | 9.28E-10 |
| DNAH17 | rs666463 | 17 | 76425480 | A | T | 0.819 | 0.189 | Imputed | 0.986 | 0.019 | 1.019 | 0.085 | 8.27E-01 | 0.829 | 0.171 | 0.071 | 1.073 | 0.023 | 1.82E-03 | 0.833 | 0.167 | 0.076 | 1.079 | 0.013 | 3.20E-09 |
| ASXL3 | rs1941685 | 18 | 31304318 | T | G | 0.522 | 0.478 | Imputed | 0.954 | 0.212 | 1.236 | 0.068 | 1.82E-03 | 0.499 | 0.499 | 0.033 | 1.033 | 0.017 | 5.73E-02 | 0.498 | 0.498 | 0.053 | 1.054 | 0.009 | 1.69E-08 |
| RIT2 | rs12456492 | 18 | 40673380 | A | G | 0.660 | 0.340 | Genotyped | 0.999 | -0.122 | 0.885 | 0.069 | 7.99E-02 | 0.674 | 0.326 | -0.104 | 0.901 | 0.018 | 4.89E-09 | 0.682 | 0.318 | -0.098 | 0.907 | 0.010 | 3.80E-23 |
| MEX3C | rs8087969 | 18 | 48683589 | T | G | 0.518 | 0.482 | Imputed | 0.962 | -0.120 | 0.887 | 0.067 | 7.34E-02 | 0.550 | 0.450 | -0.057 | 0.945 | 0.023 | 1.26E-02 | 0.550 | 0.450 | -0.058 | 0.944 | 0.010 | 1.41E-08 |
| SPPL2B | rs55818311 | 19 | 2341047 | T | C | 0.688 | 0.312 | Imputed | 0.868 | -0.034 | 0.967 | 0.076 | 6.54E-01 | 0.677 | 0.323 | -0.059 | 0.943 | 0.021 | 6.11E-03 | 0.694 | 0.306 | -0.070 | 0.932 | 0.011 | 4.18E-10 |
| CRLS1 | rs77351827 | 20 | 6006041 | T | C | 0.125 | 0.125 | Imputed | 0.979 | 0.003 | 1.003 | 0.102 | 9.76E-01 | 0.127 | 0.127 | 0.094 | 1.098 | 0.026 | 2.37E-04 | 0.128 | 0.128 | 0.080 | 1.083 | 0.014 | 8.87E-09 |
| DYRK1A | rs2248244 | 21 | 38852361 | A | G | 0.277 | 0.277 | Imputed | 0.970 | -0.048 | 0.953 | 0.077 | 5.29E-01 | 0.290 | 0.290 | 0.100 | 1.105 | 0.021 | 2.01E-06 | 0.283 | 0.283 | 0.071 | 1.074 | 0.011 | 2.74E-11 |

[a]Nalls 2019 without 23andMe data [12]
[b]Nalls 2019 with 23andMe data [12]
* SNCA rs356182 can also be found in table S4
LRRK2 G2019S (rs34637584) is occurring twice in the table, the italic being the genotyped data and the non-italic the imputed data.

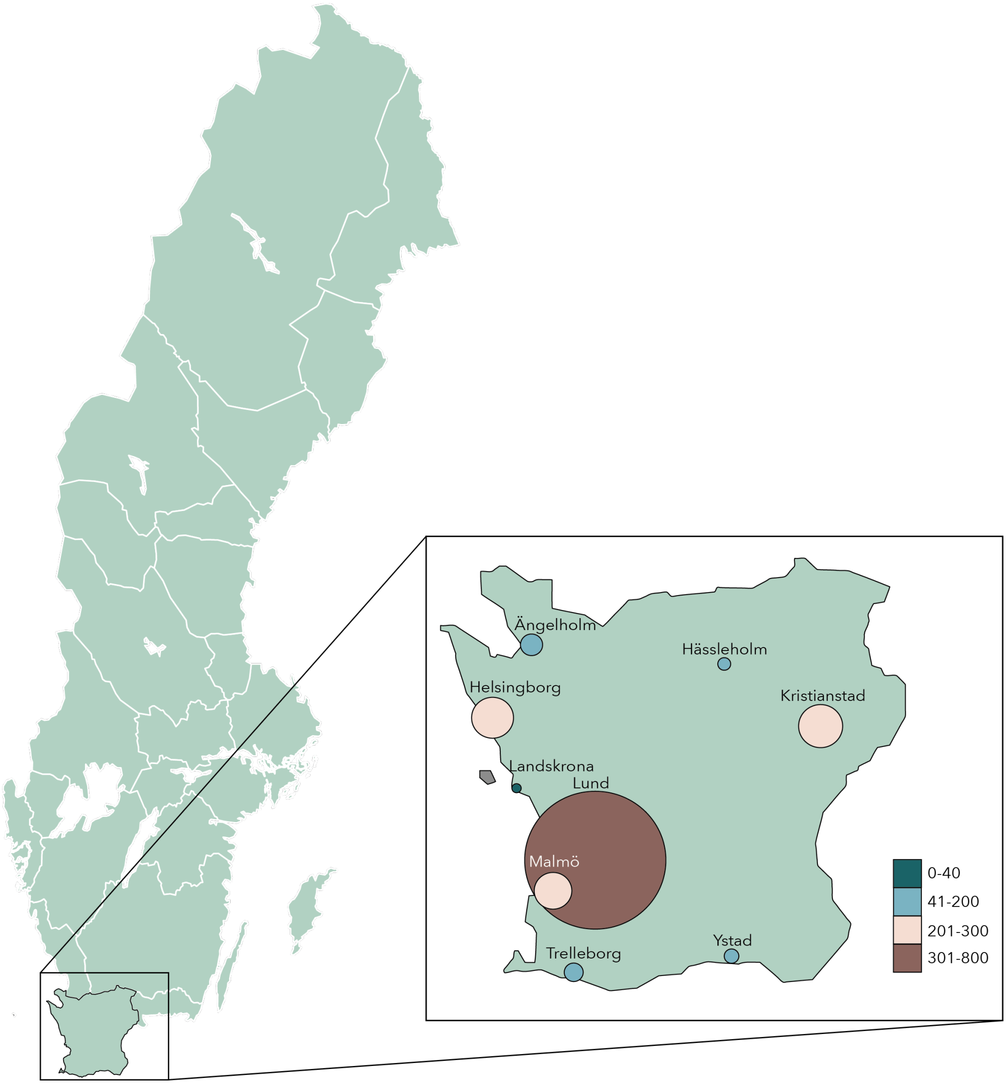


**Figure S1: Map over Sweden indicating the region of study inclusion.** The inclusion region, the southernmost province of Sweden (Scania), is enlarged. The map over Scania shows the nine different cities of study recruitment (two neurological clinics were located in Malmö). The size and color of the circles indicates the number of study participants recruited from each city.

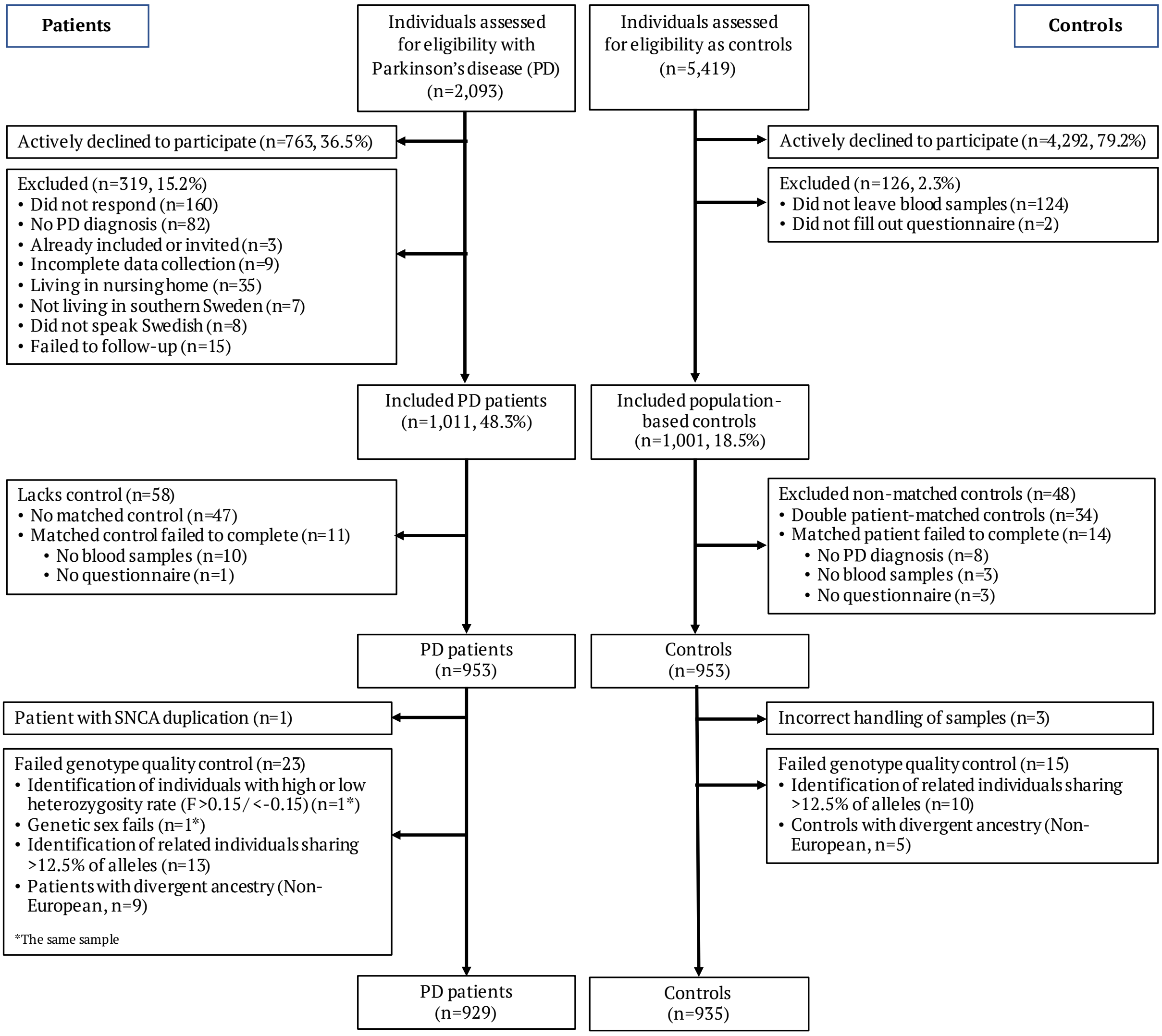


**Figure S2: Flowchart of the study participant inclusion process to MPBC**

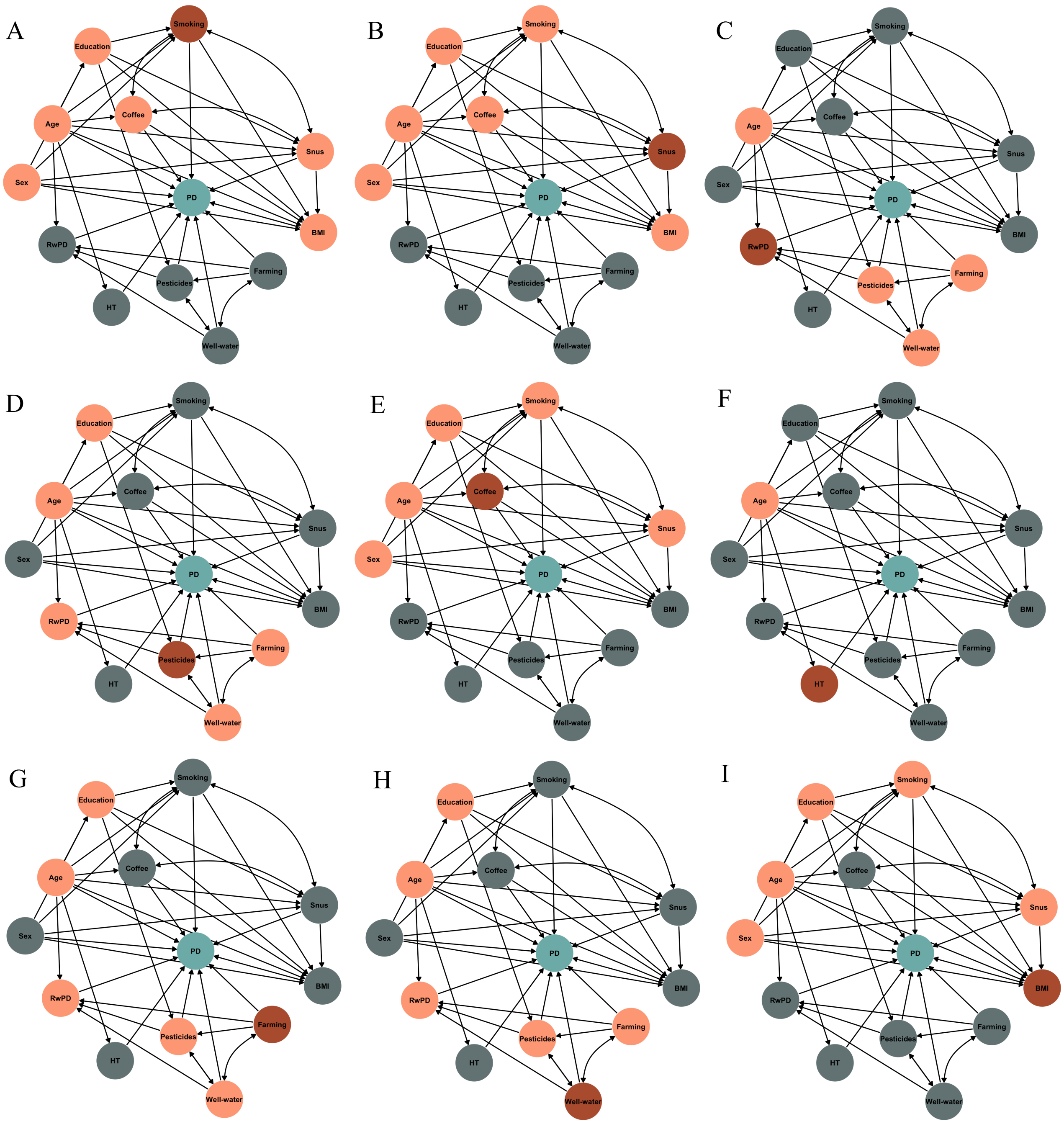


**Figure S3: Directed acyclic graphs (DAGs) for visualizing the minimal sufficient adjustment for estimating the direct effect of environmental factors on PD.**

A) Smoking, B) Snus, C) PD Heredity, D) Pesticides, E) Coffee, F) Head Trauma (HT), G) Farming, H) Well-Water, I) BMI. Red = Exposure of interest, turquoise = Outcome; PD, Orange = potential confounders to adjust for in the regression model, grey = Other variables. RwPD = Relative/s with PD, HT = Head trauma, BMI = Body mass index

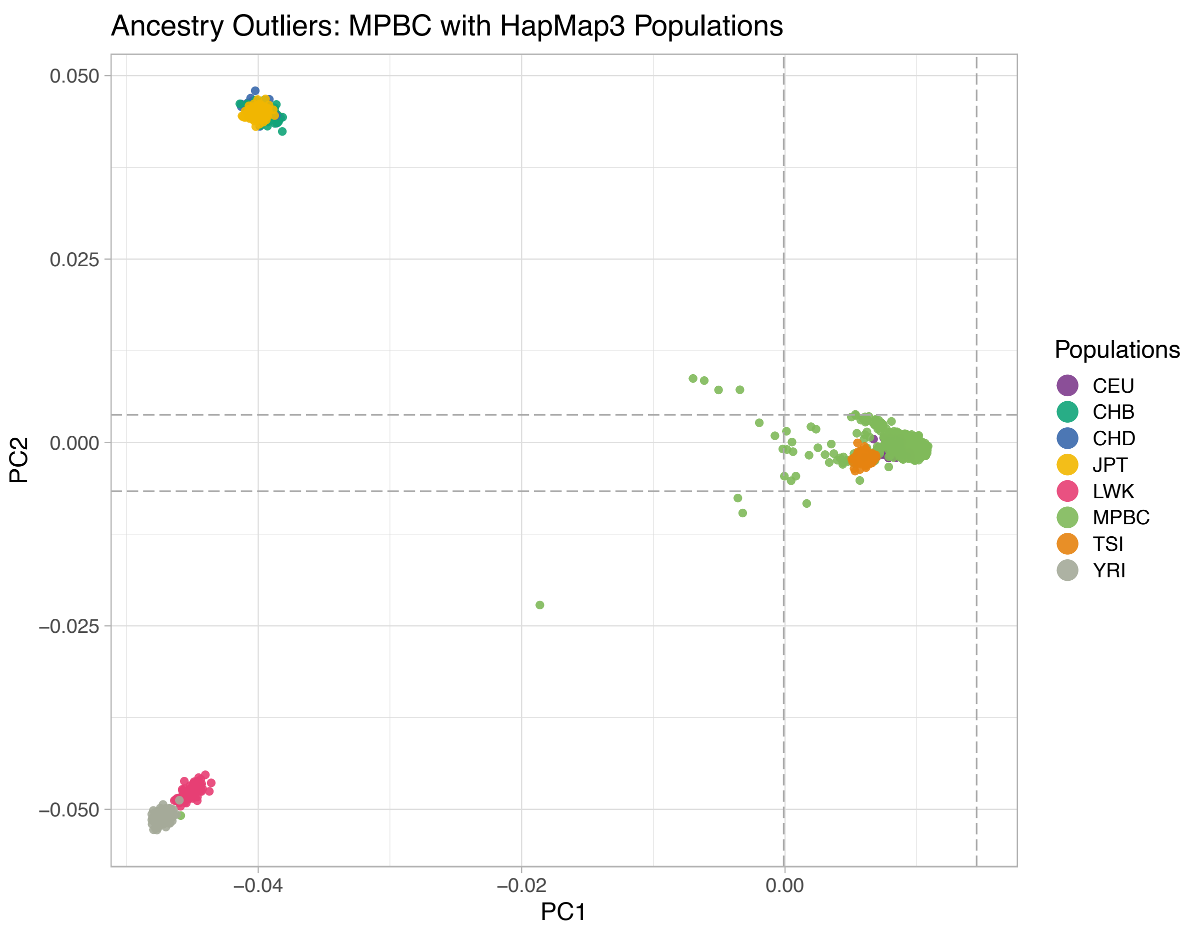


**Figure S4: Exclusion of ancestry outliers using principal component analysis (PCA).** Non-European individuals was defined as diverging >±6SD from the combined CEU/TSI population. Populations: CEU: Utah residents with Northern and Western European ancestry; CHB: Han Chinese in Beijing, China; CHD: Chinese in Metropolitan Denver, Colorado; JPT: Japanese in Tokyo, Japan; LWK: Luhya in Webuye, Kenya; TSI: Toscani in Italia; YRI: Yoruba in Ibadan, Nigeria

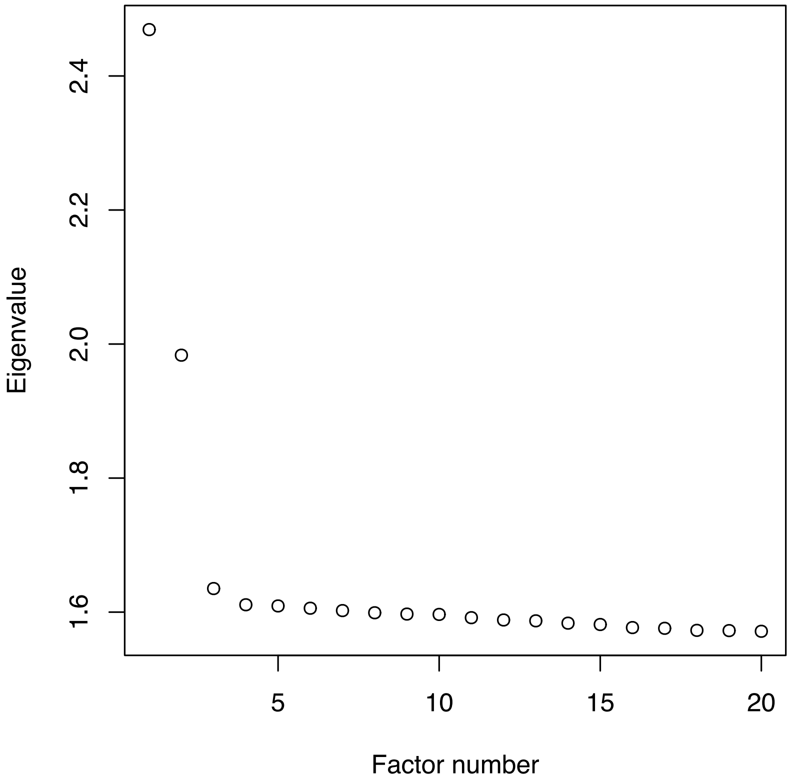


**Figure S5: Scree plot of the eigenvalues of principal components in the PCA.** Used to determine the number of PCs to add as covariates in the GWA analyses.

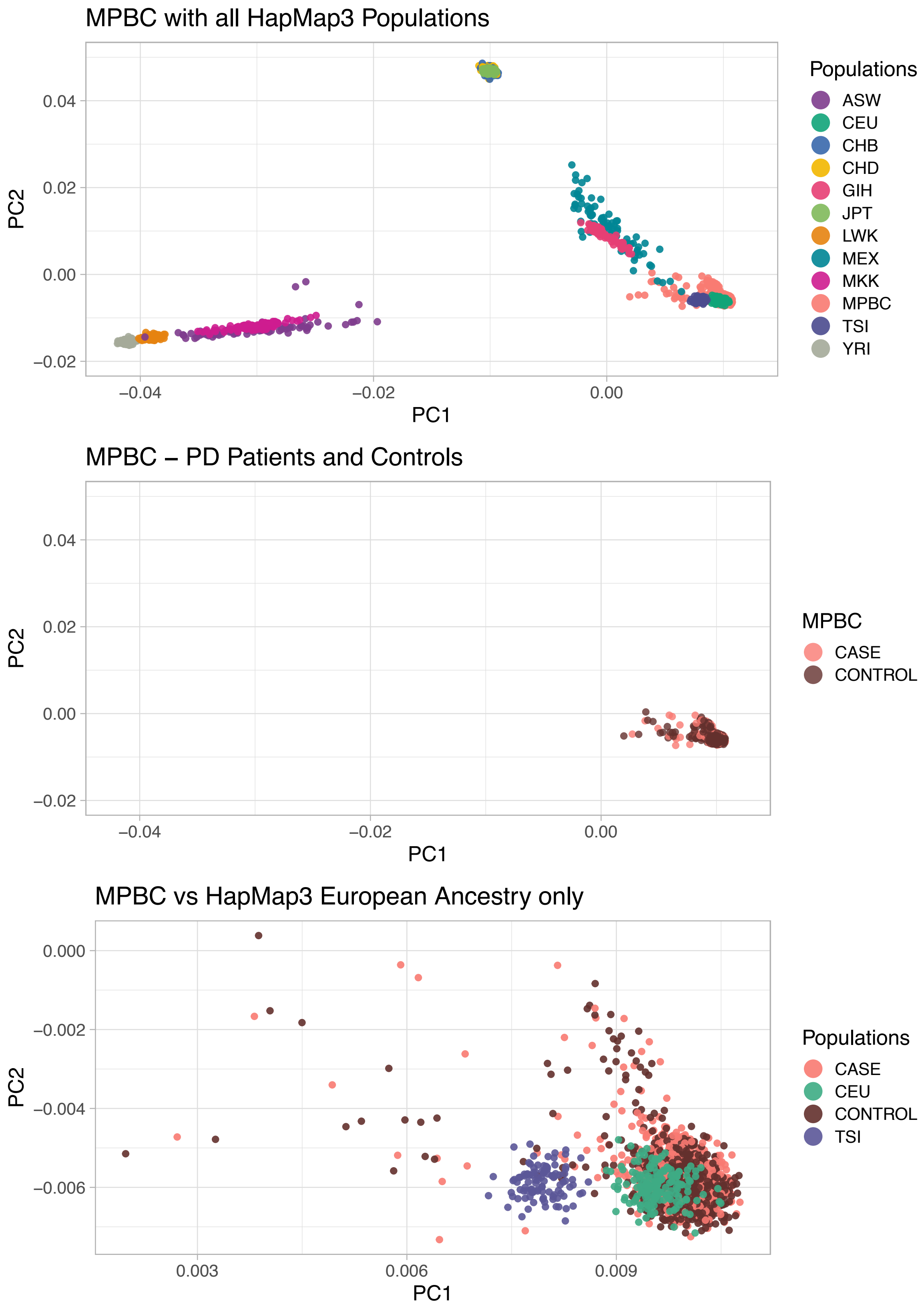
MPBC with all HapMap3 Populations
PC2
0.04
0.02
0.00
−0.02
−0.04
−0.02
0.00
PC1
Populations
ASW
CEU
CHB
CHD
GIH
JPT
LWK
MEX
MKK
MPBC
TSI
YRI
MPBC – PD Patients and Controls
PC2
0.04
0.02
0.00
−0.02
−0.04
−0.02
0.00
PC1
MPBC
CASE
CONTROL
MPBC vs HapMap3 European Ancestry only
PC2
0.000
−0.002
−0.004
−0.006
0.003
0.006
0.009
PC1
Populations
CASE
CEU
CONTROL
TSI

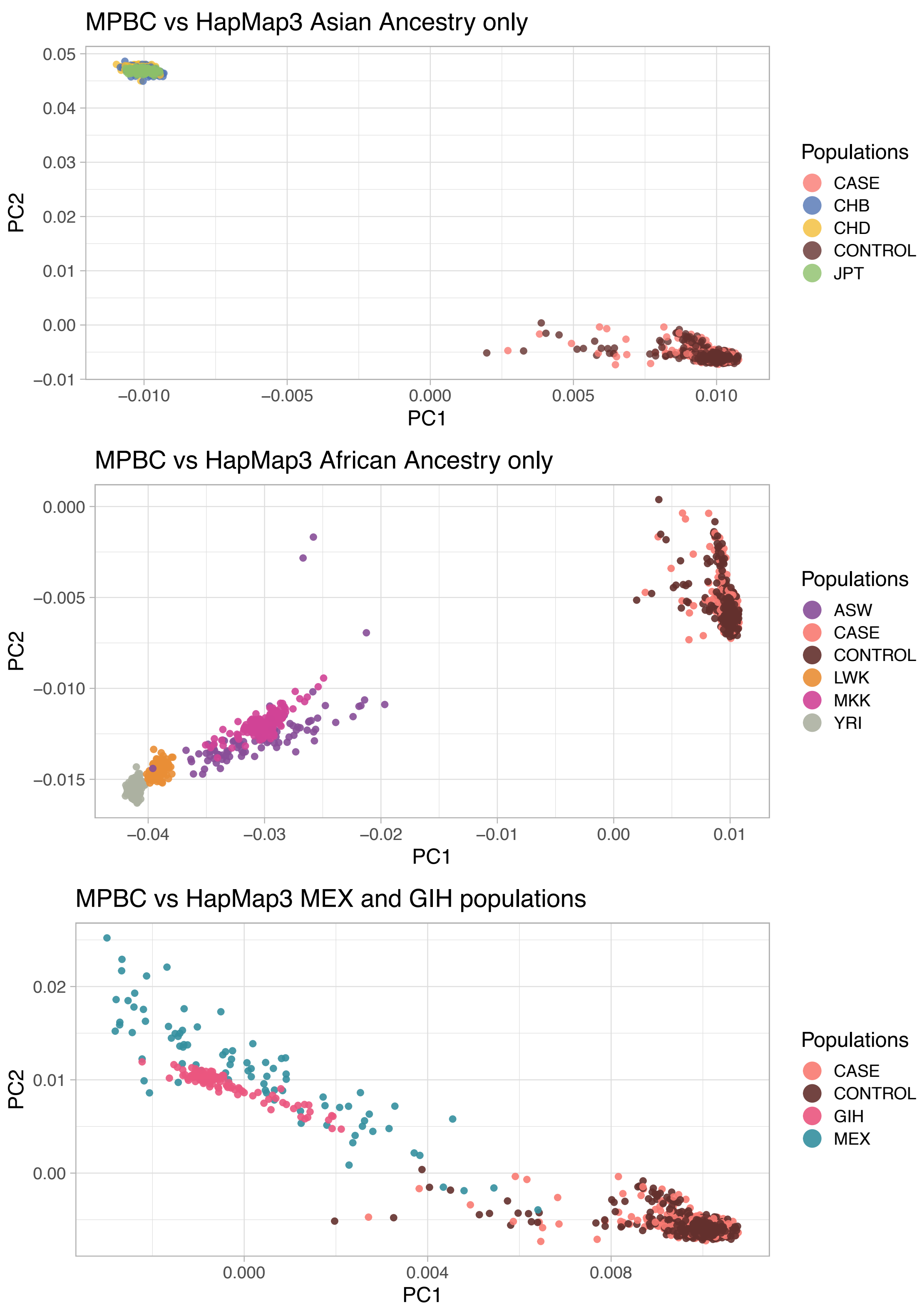


**Figure S6: Swedish cohort MPBC population stratification with HapMap3 populations using PCA.** Populations: ASW: African ancestry in Southwest USA; CEU: Utah residents with Northern and Western European ancestry; CHB: Han Chinese in Beijing, China; CHD: Chinese in Metropolitan Denver, Colorado; GIH: Gujarati Indians in Houston, Texas; JPT: Japanese in Tokyo, Japan; LWK: Luhya in Webuye, Kenya; MXL: Mexican ancestry in Los Angeles, California; MKK: Maasai in Kinyawa, Kenya; TSI: Toscani in Italia; YRI: Yoruba in Ibadan, Nigeria

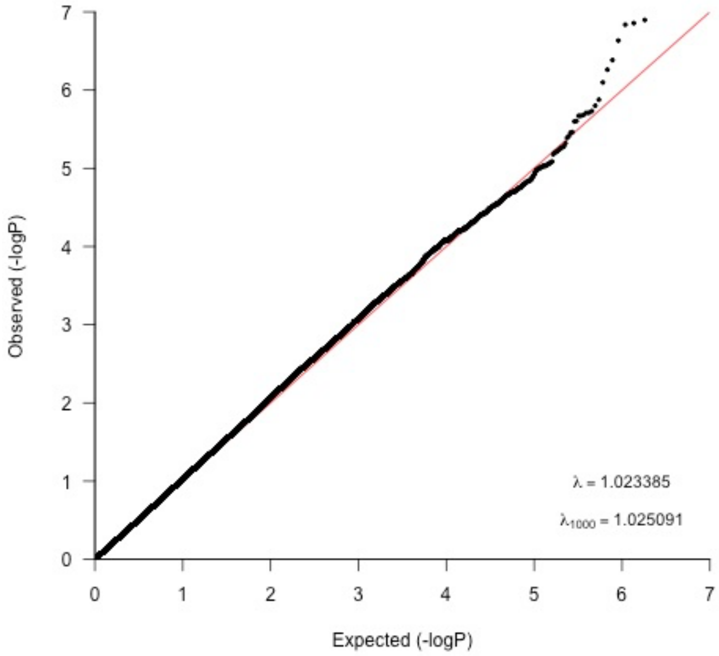


**<u>Figure S7: Quantile-quantile-plot for PD GWAS with a total of 5,445,841 SNPs (MAF > 5%) tested for 929 PD patients vs 935 controls.</u>**

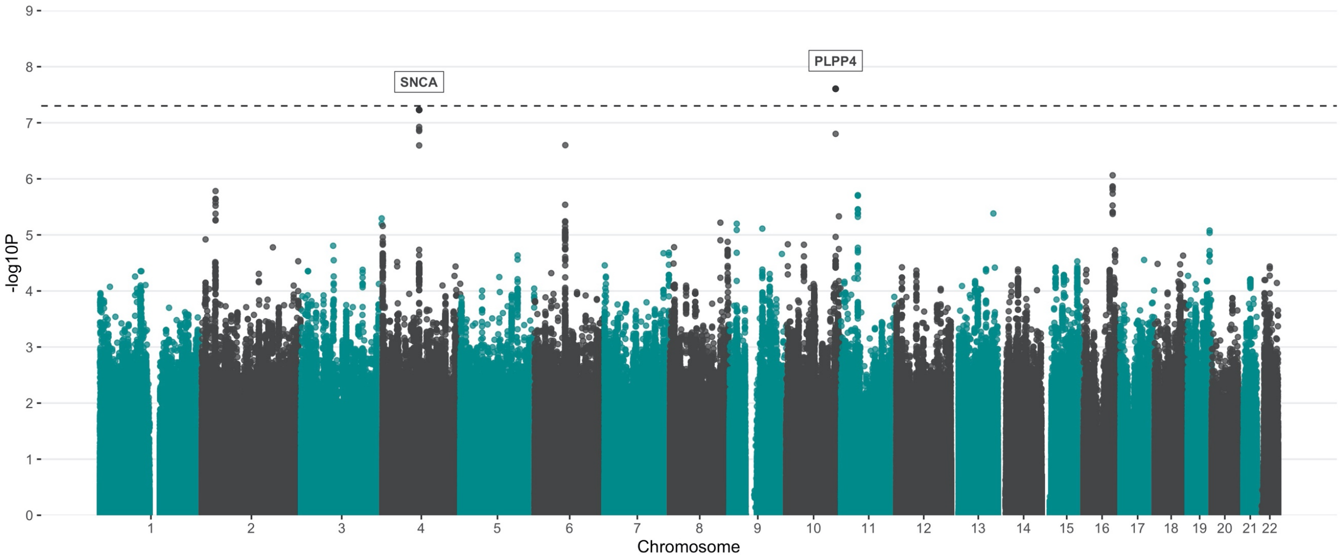


**Figure S8: Manhattan plot showing the result from PD GWA analysis following imputation with the TOPMed Imputation Reference panel.** A total of 6,214,098 variants were included in the analysis following post-imputation QC (MAF > 5%, Rsq >0.3).

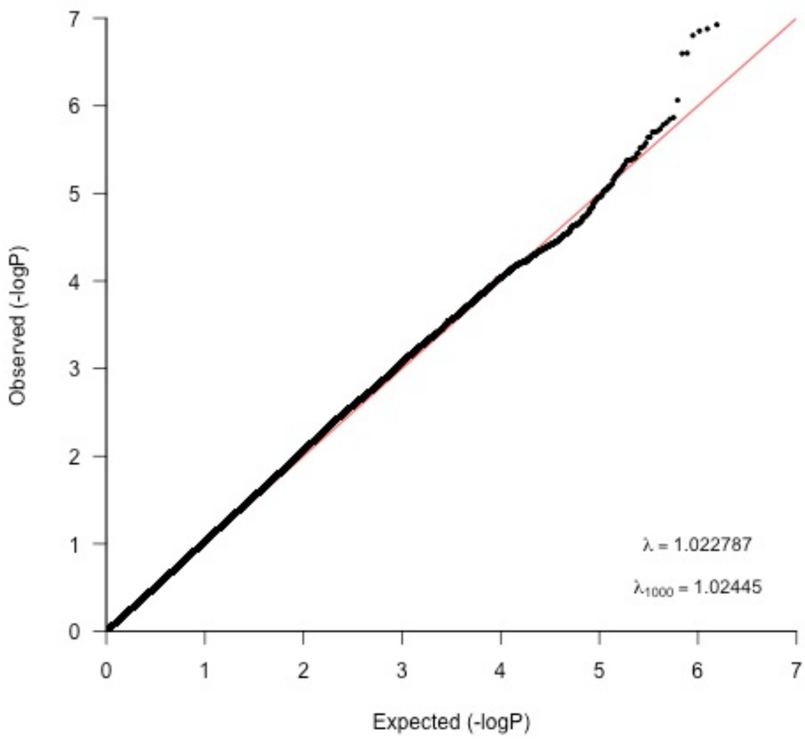


**Figure S9: Quantile-quantile-plot for PD GWAS following imputation with the TOPMed Imputation Reference panel and post-imputation QC (MAF > 5%, Rsq > 0.3)**

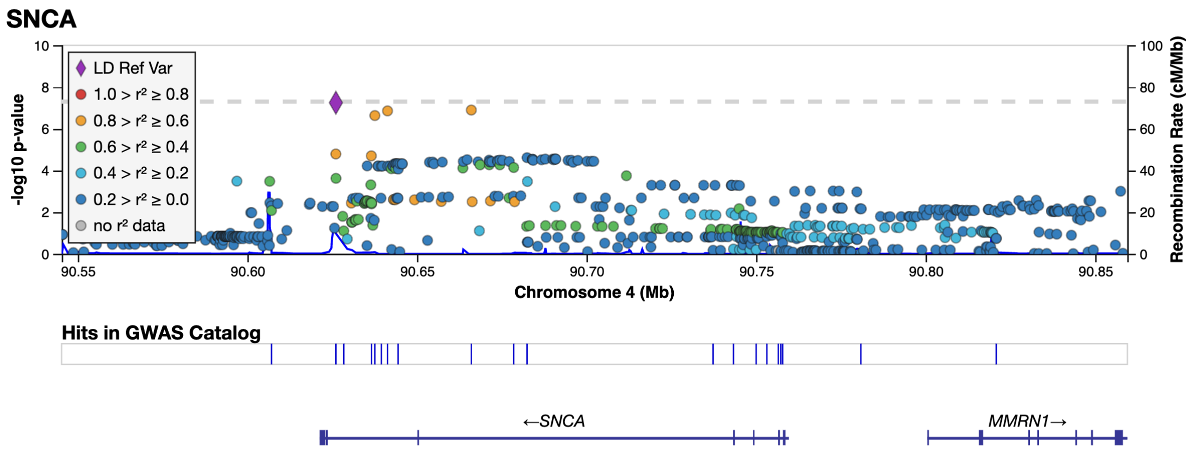


**Figure S10: LocusZoom plot for PD GWAS *SNCA* loci.** Imputed and genotyped variants passing QC in the *SNCA* gene +/- 100 kb (chr4: 90545250 - 90859466) mapped to genome build GRCh37. The variant with lowest p-value (index) is indicated as a purple diamond. Marker colors indicate the strength of LD as r^2 between the index variant and other variants in the 1000 Genomes EUR population.

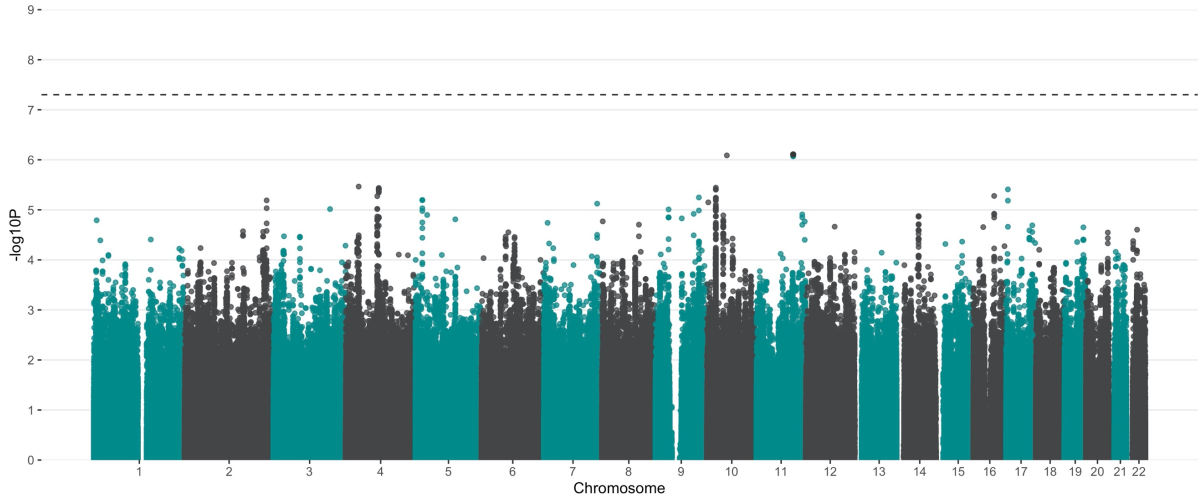


**Figure S11: Manhattan plot showing the results from the PD age at diagnosis (AAD) GWAS.** Data for AAD was available for 792 of 929 PD patients (85.3%) in the cohort and the analysis was adjusted for sex and PC1-5. Analysis was run using 5,440,801 variants following exclusion of variants with a MAF <5% in the group.

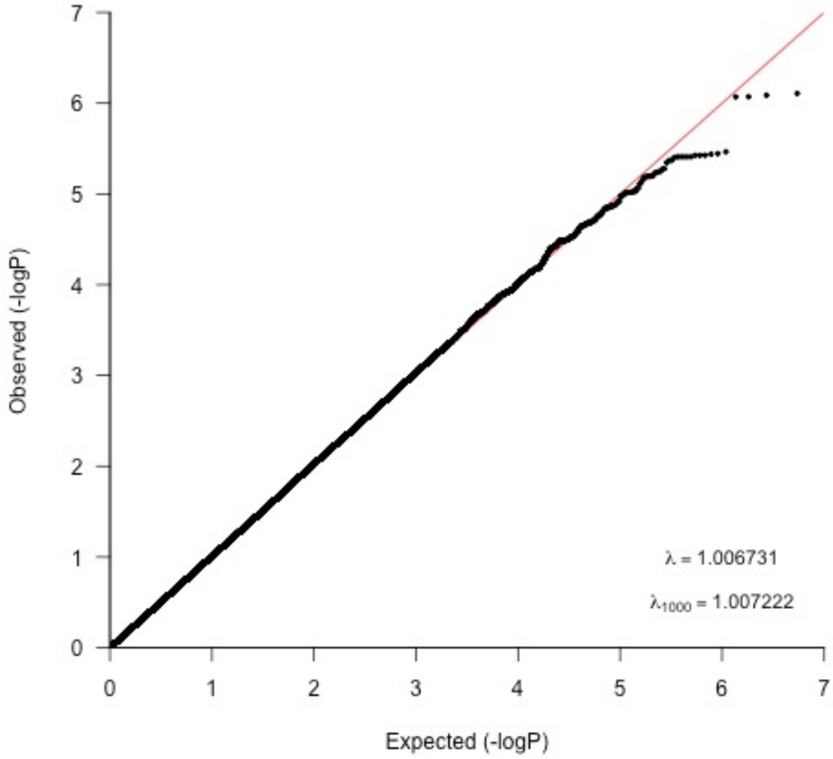


**Figure S12: Quantile-quantile-plot for the age at PD diagnosis GWAS.**

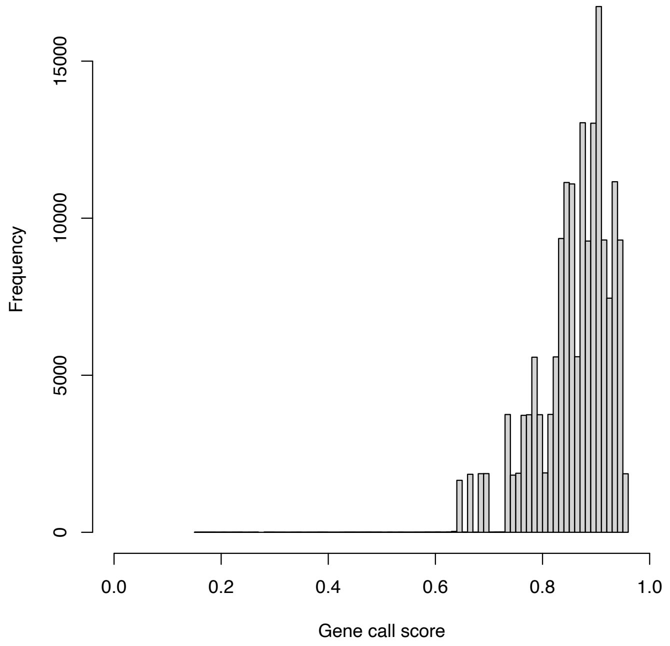


**Figure S13: GenCall scores for genotyped variants (n=92) in *PLPP4* ±100 kb**

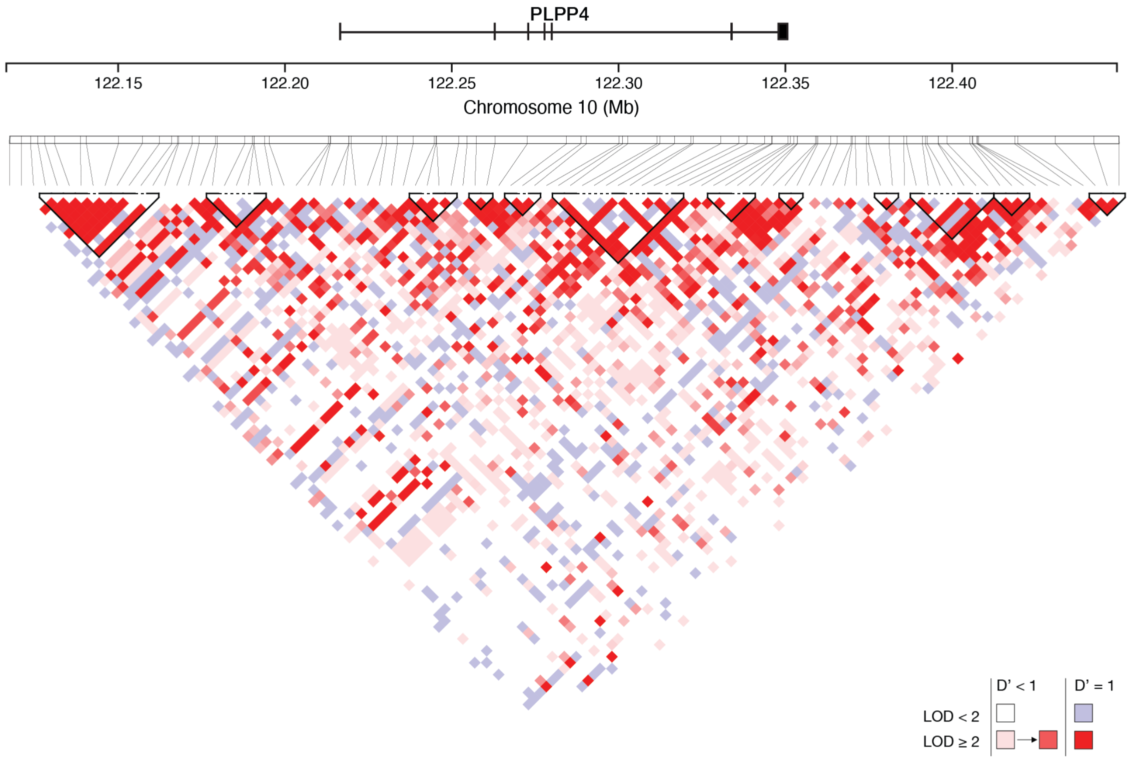


**Figure S14: Linkage disequilibrium (LD) plot in the PLPP4 locus.** LD heatmap showing the LD (D') between the genotyped variants in the region in the MPBC cohort. Note that the location of variants in the D heatmap can be shifted relative to the chromosomal position.
